# Co-design and refinement of an optimised antenatal education session to better inform women and prepare them for labour and birth

**DOI:** 10.1101/2023.12.22.23300473

**Authors:** Abi Merriel, Miriam Toolan, Mary Lynch, Gemma Clayton, Andrew Demetri, Lucy Willis, Narendra Mampitiya, Alice Clarke, Katherine Birchenall, Chloe de Souza, Emma Harvey, Tamarind Russell-Webster, Eva Larkai, Mariusz Grzeda, Kate Rawling, Sonia Barnfield, Maggie Smith, Rachel Plachcinski, Christy Burden, Abigail Fraser, Michael Larkin, Anna Davies

## Abstract

**Objective:** To co-design, implement, evaluate acceptability and refine an optimised antenatal education session to improve birth preparedness.

**Design:** There were four distinct phases: co-design (focus groups and co-design workshops with parents and staff); implementation of intervention; evaluation (interviews, questionnaires, structured feedback forms), and systematic refinement.

**Setting:** A single maternity unit with approximately 5,500 births annually.

**Participants:** Postnatal and antenatal women/birthing people, birth partners and clinicians. ***Outcome measures:*** To establish whether the optimised session is deliverable, acceptable, meets the needs of women/birthing people and partners, and refine it with input from parents, clinicians and researchers.

**Results:** The co-design was undertaken by 35 women, partners and clinicians. Five midwives were trained and delivered 19 Antenatal education (ACE) sessions to 142 women and 94 partners. 121 women and 33 birth partners completed the feedback questionnaire. Women/birthing people(79%) and birth partners(82%) felt more prepared after the class with most participants finding the content very helpful or helpful. Women/birthing people perceived classes were more useful and engaging than their partners. Interviews with 21 parents, a midwife focus group and a structured feedback form resulted in 38 recommended changes: 22 by parents, 5 by midwives and 11 by both. Suggested changes have been incorporated in the training resources to achieve an optimised intervention.

**Conclusions:** Engaging stakeholders (women and staff) in co-designing an evidence informed curriculum resulted in an antenatal class designed to improve preparedness for birth, including assisted birth, that is acceptable to women and their birthing partners, and has been refined to address feedback and is deliverable within NHS resource constraints. A nationally-mandated antenatal education curriculum is needed to ensure parents receive high-quality antenatal education that targets birth preparedness.

**Key messages:** *What is already known:* Antenatal education is used to prepare women/birthing parents for labour, birth and the postnatal period, but it has been eroded. Antenatal education has potential to support women/birthing parents in developing their expectations around labour and the postnatal period, via improved health literacy. Improving antenatal education could be impactful as the expectation-experience gap is linked to post traumatic stress disorder.

*What this study adds:* We have shown that a co-designed, optimised antenatal class can provide information helpful to parents and important to staff, within the constraints of the NHS resources

*How this study might affect research, practice or policy:* This study can be used to understand what parents need from antenatal education, and how to begin to address the expectation-experience gap.

## Introduction

Antenatal education(ANE) has been used to prepare women/pregnant people for labour, birth for many years.^1^ It is a vital element of antenatal care and is incorporated into the NICE guidelines.^2^ ANE contributes to practical preparation, but it can contribute to a woman/pregnant person’s expectations and experience of labour and birth and consequently their psychosocial outcomes.^3^

When considering what is important to them about birth, women prioritise the physiological birth of a healthy baby. However, when things do not go according to their plan, they wish to retain a sense of personal achievement and control through active decision-making.^4^ Empowering women/pregnant people to participate in this process through high-quality antenatal education has been shown to mediate childbirth satisfaction.^5^

Antenatal preparation has the potential to support women/pregnant people in developing their expectations. This is important because an expectation-experience gap increases risk of post-traumatic stress disorder (PTSD).^5^ The origins of a woman/pregnant person’s PTSD are not likely to lie solely with birth,^6^ but high rates of stress-related symptoms are experienced following unanticipated intervention in labour. Up to half have PTSD two months after unplanned Caesarean compared to 24% at six weeks after vaginal birth.^7^ Risk factors for PTSD include subjective birth experience relating to negative emotions and lack of control or agency, operative birth and lack of support from staff during birth.^6^ Up to 1.5% of women experience PTSD six months postnatally.^8^ There may also be a link between birth expectations and depression.^5^ Good quality ANE provides an understanding of common interventions that might become necessary, and could attenuate the expectation- experience gap.

ANE provision is variable, less than a third of women are offered antenatal classes.^9^ ANE is available within the NHS, privately-for-profit or not-for-profit, by clinicians and allied healthcare staff, or by trained antenatal educators. ANE can be traditional information provision classes or focussed on self-directed coping strategies e.g. hypnobirthing; some pregnancy exercise classes also provide elements of education and preparation. Different classes may have a particular focus (for example physiological birth) or a clear goal to provide evidence-based information. This area is un-regulated and although NICE recommend ANE for all women in their first pregnancy, they do not provide comprehensive guidelines on what should be covered in classes.^2^

Prior to this intervention development study, we conducted focus groups with 48 postnatal women.^10 11^ They described limited discussion of common interventions (e.g. assisted birth, induction) and birth experiences (e.g. perineal trauma) resulting in them believing that these events were infrequent and unnecessary to learn about. Participants reflected that receiving sensitively provided information about the frequency and nature of interventions and common events during birth was important and could support psychological health if birth experiences were not as expected. When discussing ANE with midwives (unpublished) we found many were not provided with specific training to deliver ANE, nor did they enjoy or want to deliver it. Furthermore, midwives often designed the class materials themselves, with little assistance or guidance.

We aimed to improve antenatal education provision by co-designing and refining an optimised antenatal education package (ACE).

## Methods

### Ethical approvals

This study was approved by Wales Research Ethics Committee 6 (reference: 21/WA/0091).

The methods are presented in the four phases: Co-design (2019), trial implementation (2021-2022), evaluation and refinement (2021-2022).

### Research team and organisational commitment

The initial research team was made up of obstetricians (AM, MT, SB, CB), midwives (MLy, MS), psychologists (ADa, EA, MLa), service users (KR), NCT representative (RP), epidemiologist (AF), project manager and medical students (TRW,EL). As the project progressed and was impacted by COVID-19 additional trainee obstetricians (ADe, CdS, KB), doctor-in-training (AC) and medical students (LW, NM, EH) joined the research team. ACE was supported by the Head of Community Midwifery (MS) and the then Head of Obstetrics (SB). This facilitated delivery of the study and will enable future roll out.

### Patient and public involvement

Parents were involved from the inception of the study and played an active role in the design. A parent sat on the study steering group (KR) and parents co-designed the ACE intervention, drawing on the data generated in focus groups/survey studies.

### Study setting and context

This study was delivered in a single hospital in the South-West of England with 5500 births annually. It was conducted before and during the COVID-19 pandemic (2019 to 2022) when there was limited provision of ANE and limited contact between staff and women.^12^

### Frameworks to inform intervention development

We used the Medical Research Council’s Complex Intervention Development Framework (2008) to develop the ACE intervention.^13^ The consolidated framework for advancing implementation science (CFIR) was used to plan and support initial testing and further development of the intervention,^14^ to ensure preparedness for implementation.

## Phase 1: Co-design

Co-design was chosen as a method to involve service users in the development of the ACE session because patient experience, outcomes and safety are linked.^15^ We saw the best way to improve patient experience as being to involving service users in the intervention design. We used and adapted experience-based co-design (EBCD)approach.^16 17^ Figure 1 shows the EBCD steps and our adaptations to them. We included the two core co-design elements: service user experience data and including service users in the design.^17^ The planned output from the co-design was a two-hour ACE session on labour and birth, and materials to deliver the session.

**Figure 1:**
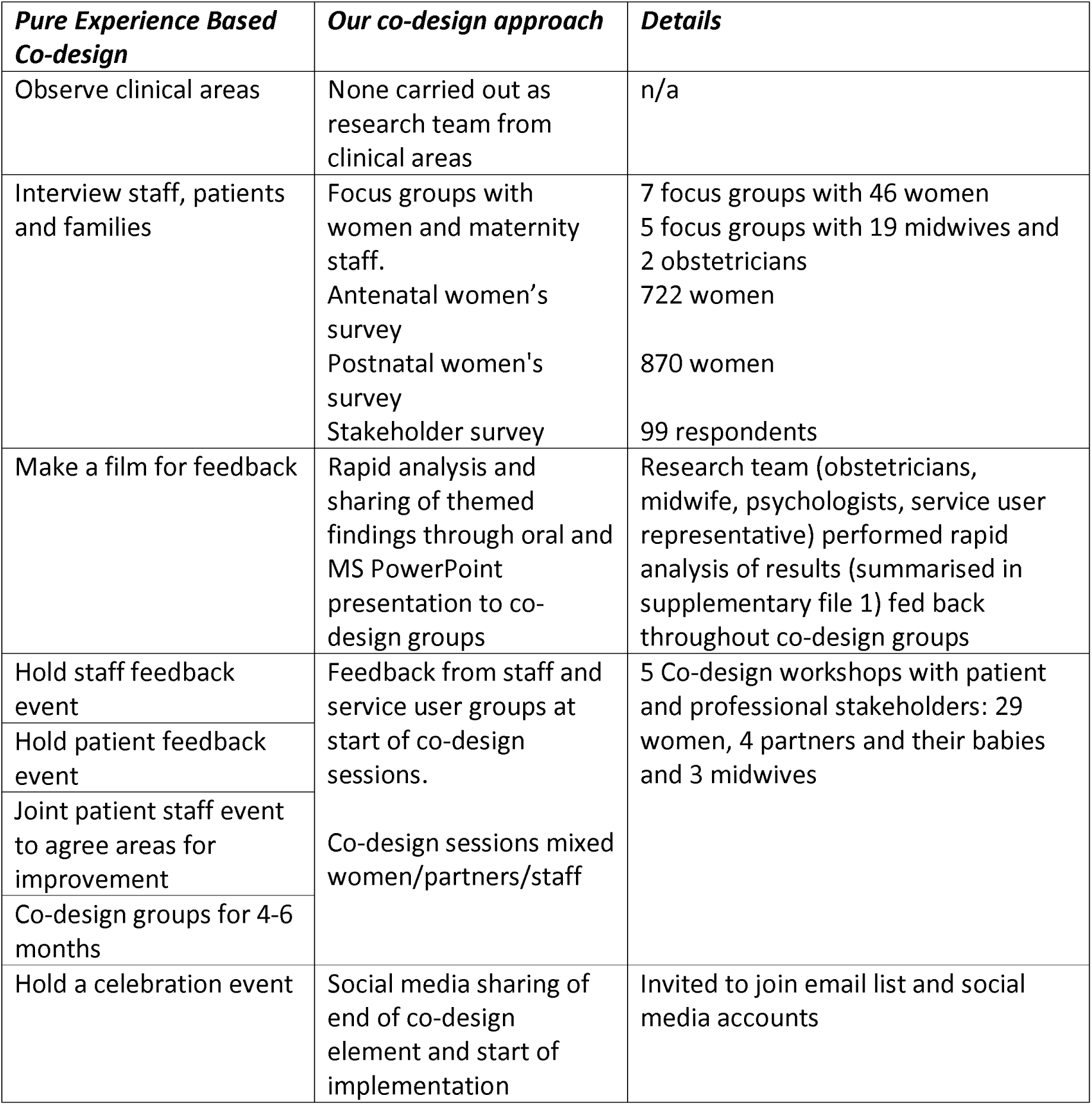
Our modifications to Experienced based co-design.

## Phase 2: Implementation

### Recruitment of antenatal educators

We recruited community midwives who deliver antenatal education. Those agreeing to take part were invited to a training session. All time contributed to training and delivery of ACE was remunerated via the midwifery ‘bank’ system.

### Antenatal education group attendees

We invited women/birthing people who were over 18 years of age, more than 24 weeks pregnant and in the care of the local trust to participate. We informed them that the ACE session was a supplement to their NHS offered session. They were asked to invite their partners if they wished. Recruitment was via 1) social media, 2) community midwifery referrals, 3) telephone calls to all eligible pregnant women/birthing people. Women/birthing people and their partners were given written information via email. They then booked into a session on the date of their choice, completed an online consent form and provided basic demographic information.

### Impact of the COVID-19 pandemic

We paused this study as face-to-face ANE was halted soon after the co-design groups had finished. We re-started this study when face-to-face interactions were permitted, and ran ACE classes in a COVID-secure way. Parents were therefore invited to a central location in the evenings, rather than the originally planned local venues.

### Training in delivering the ACE session

A training session was held for midwives. As provision expanded, midwives new to the programme attended an ACE session delivered by an experienced facilitator and discussed it with her.

### Cost of implementation

Reusable resources included a projector, printed posters and attachable reusable stickers, costing £150-200. Each woman/birthing person requires a printout of the ‘what is important to me’ tool. Existing resources (e.g. pelvis and doll, kiwi cups) were drawn upon. After midwifery training, the remainder of the costs were identical to existing costs to deliver antenatal education.

## Delivery

The session was 2 hours long and (due to COVID-19) restricted to 10 participants and their partners. The research team observed each session, noting attendance and contemporaneously recording the extent to which the intervention was delivered as intended (fidelity), and the session length. Women received a £10 voucher for their expenses.

## Phase 3: Evaluation

The ACE class was evaluated/refined in 5 ways:

1. Immediate debrief with midwife delivering the session This discussion between the midwife and research team was recorded on a structured form to identify successes and areas for improvement. Feedback was reviewed after each session by the study lead(AM)/trial manager(ADa) to identify any immediate changes to implement.
2. Online questionnaire for women/birthing people and birth partners. An online survey hosted in REDCap^18^ was sent out within 2 weeks of attending the session. Participants were asked to rate the content, delivery and resources and how useful the session was in preparing them for childbirth. They were asked if they had attended other ANE, and if so to compare the ACE sessions to these. Quantitative data were analysed in STATA, and themes from free-text feedback were coded by a researcher. This was analysed at the end of the study to provide overall feedback and identify additional refinements.
3. Semi-structured feedback interviews with women/birthing people +/- birth partner approximately 2 weeks after attending the ACE session and/or 4-6 weeks after birth. At the end of the classes women/birthing people +/- their birth partners were asked to express interest in being interviewed about the class. Interviews were conducted either before their baby was born (approximately 2 weeks post ACE Class), or 4-6 weeks after their baby was born. Parents could take part in one or both interviews. These could be conducted alone or with their partner in attendance. We purposively sampled parents who had attended sessions delivered by different midwives, both at the start and end the implementation capture a range of experiences. Interested parents were offered written information and a mutually convenient time for a telephone interview. Informed consent was obtained via a form hosted in REDCap.^18^ The interviews were audio recorded and rapid thematic analysis performed directly from the audio recordings to identify areas for change and areas where feedback was positive. A £10 voucher was provided.
4. Midwife focus group We undertook a single one-hour focus group the midwives who delivered the ACE class to gather feedback about their experiences of the class, the manual, and any required improvements. The group was audio recorded and rapid thematic analysis carried out from the audio files. A £10 voucher was provided.

## Phase 4

### Refinement and reporting of findings

The findings from the immediate session feedback were iteratively implemented into the intervention. The findings from the online questionnaires and from the focus groups were analysed once at the end of the study and therefore refinements were only included in the updated ACE resources (supplementary file 2).

Potential changes to the manual and materials, were recorded in an adapted Table of Changes.^19^ This shows the changes suggested, how frequently, whether it was feasible and reasonable to make the change and what change was actually made. This provided a systematic, rigorous approach to identifying potential changes, facilitating discussions between the study team, and agreeing final changes.

## Results

### Phase 1: Co-design

Five co-design groups were undertaken with 29 women, 4 partners, 3 maternity staff. They developed the concept for the ACE class, the topics, planned the materials, and discussed the importance of training for the staff.

The group designed the ACE class around a river journey, conceptualising the process of birth as a journey down the river, with a winding course that represented the different stages of labour that may be experienced before reaching birth. The river served as a metaphor to illustrate that labour and birth could take a number of different courses, but all would end in a postnatal bay with their baby, where support from friends, family and healthcare professionals would be available.

The topics identified in response to the focus group data were: differing birth journeys (spontaneous vaginal, induction, assisted vaginal birth, planned caesarean, unplanned caesarean); coping with labour and birth (pharmacological and non-pharmacological); the immediate postnatal period; birth preferences; and social support. The co-design group believed a variety of different birth experiences should be reflected within the ACE session. We therefore made videos of their experiences of vaginal births, assisted vaginal births, quick and long inductions of labour and planned and emergency caesarean births.

The co-design group planned a ‘what is important to me’ preferences tool, to support attendees in considering their birth preferences. The tool focuses on the birth of a healthy baby at the end of labour and how their preferences could aim to achieve this(supplementary file 2).

### Phase 2: Implementation

A 3-hour training session for midwives was designed and delivered online and in person as desired. The ACE manual provided a session outline. Five midwives ran 19 ACE sessions, delivered to 142 women and 94 partners. On two occasions midwives were unable to attend within 6-12 hours of the session, the project lead (AM – obstetrician) delivered the sessions to avoid inconvenience. The demographics of those attending the session are presented in table 1.

**Table 1:**
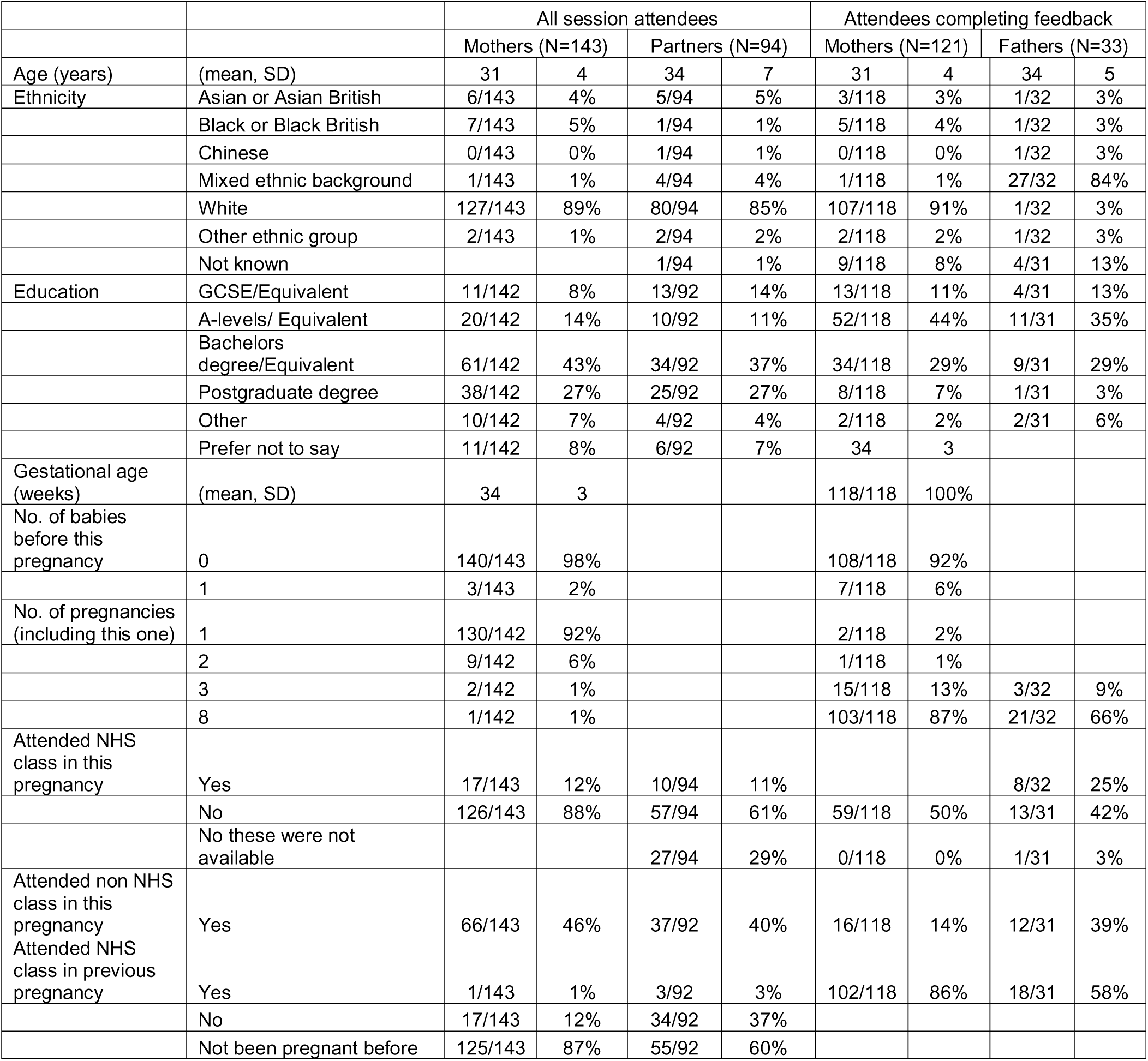
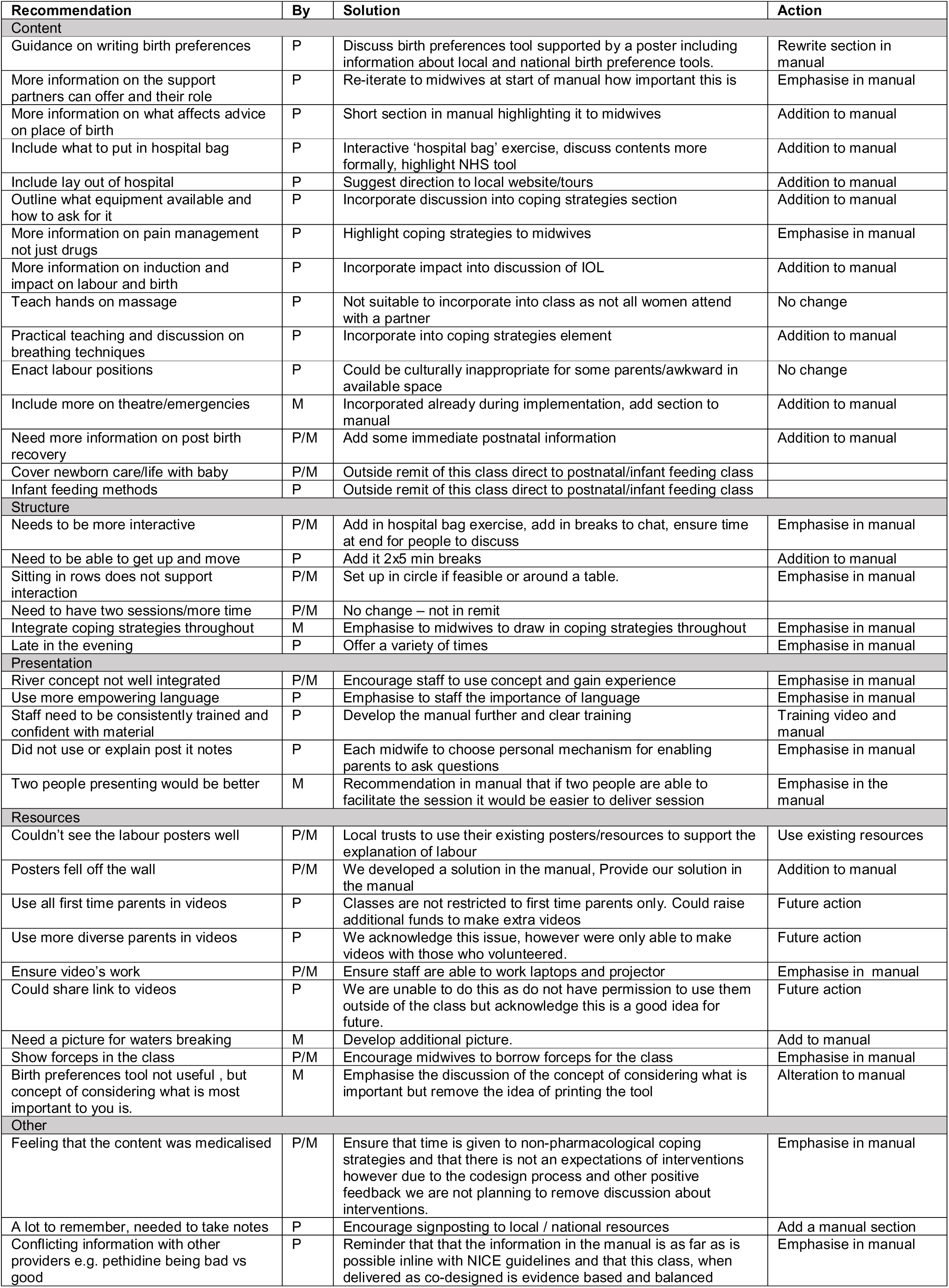
Demographics of women and birth partners attending the sessions and completing feedback.

### Phase 3: Evaluation

#### Fidelity of delivery

During the 19 sessions, most of the class content was covered consistently; in all or a majority of sessions the following content was addressed or partially addressed: birth journeys (n= 19/19), coping strategies (n= 19/19), social support (n=14/19), partner support (n=18/19). A social opportunity was provided inconsistently (n=8/19). Across groups, materials and resources were well utilised with the exception of the ‘what is important to me’ birth preferences tool which was not used in over half of the classes (n=8), although it was explained in the majority (n=17/19). Supplementary file 3 provides detailed observations and feedback.

#### Feedback from participants Interviews

21 Interviews were conducted: 13 antenatally, 7 postnatally. One couple completed both antenatal and postnatal interviews. For 3 interviews, women were interviewed with their partners. Participants valued attending a class with an NHS professional, feeling that it was an opportunity to get good-quality information about their local setting and the care they could expect. Most reported that the river concept was useful and that while a great deal of information was being given, it met their needs and was not overwhelming. Several women felt that more information around the impact of decisions, for example induction of labour, would be beneficial. They desired more opportunity for social interaction between class participants. Supplementary file 4 contains a summary of themes and supporting quotes.

#### Questionnaires

121 women(W) and 33 birth partners(BPs) completed the online feedback form following the class (demographics in table 1). Most participants attended in their third trimester, were aged 30-35, were of white ethnicity(89% W, 85% BPs) and were university educated (70% W, 64% BPs).

When considering how prepared the participants felt after the class, 79% of women and 82% of birth partners felt more prepared than they were beforehand. The majority reported having improved knowledge of strategies to cope with pain (W 79%, BP 82%) and to use if things didn’t go according to plan (W 81%, BP 85%). Further feedback is provided in figure 2. Women found the sessions more useful and engaging than their birth partners. However, when considering the specific areas of information provided (figure 3) most women and their partners found much of the session content to be very helpful or helpful.

**Figure 3:**
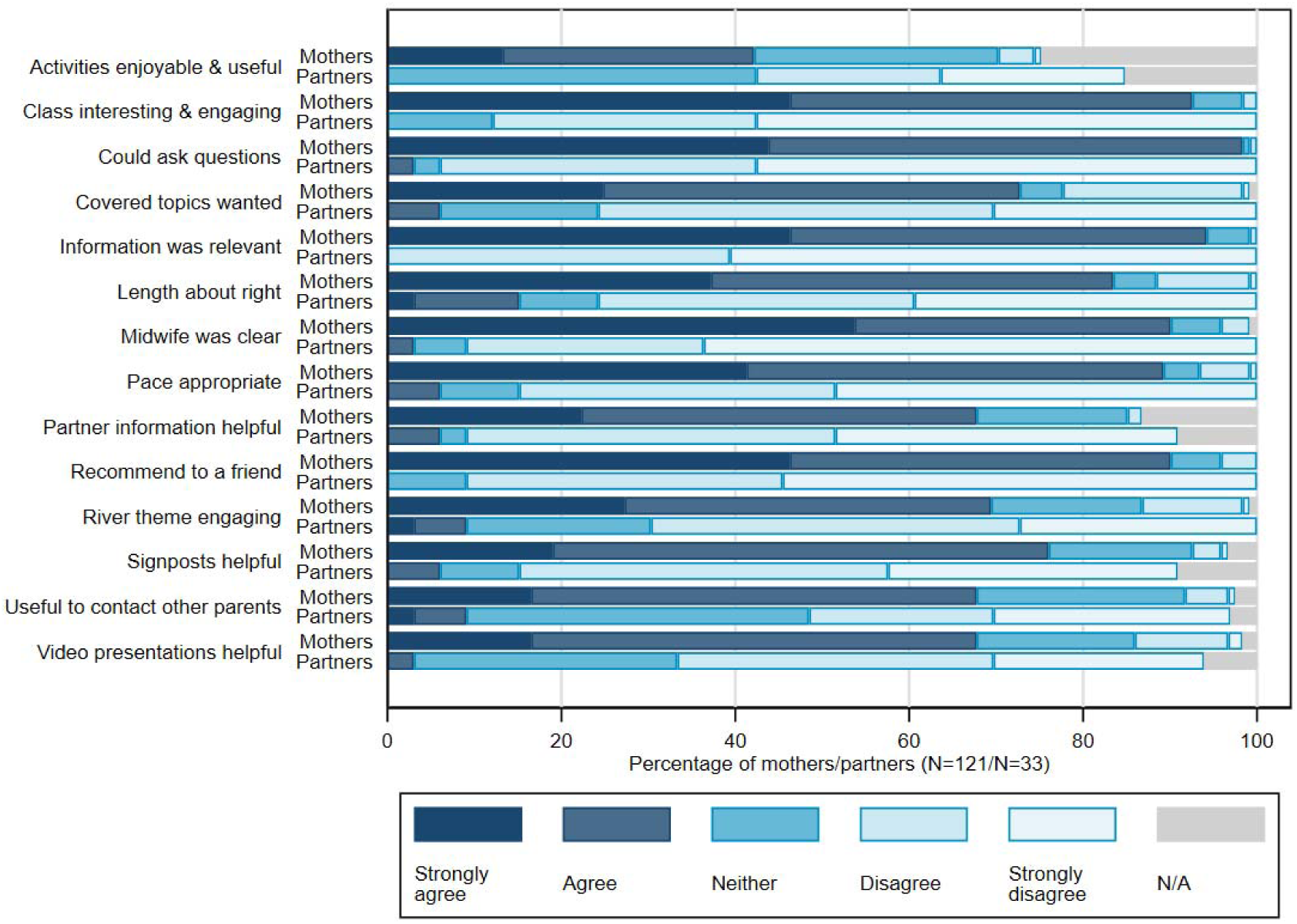
Feedback by Women and Birth partners.

**Figure 4:**
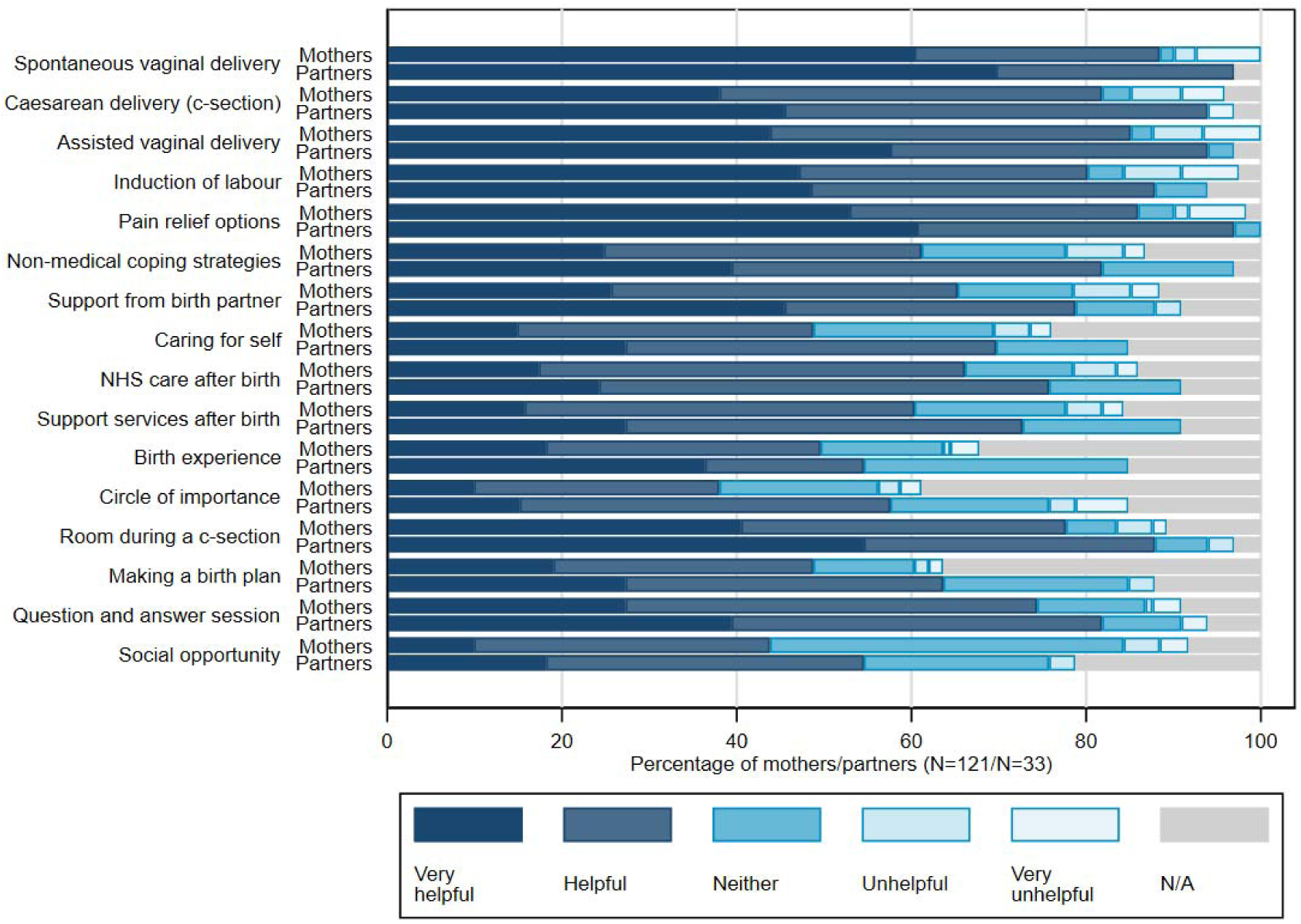
Usefulness of class according to women and birth partners.

121 women responded to questions about the attendance of the birth partner, of these 90 (74%) women reported that their birth partners attended the class. Of those whose partner did not attend, 21 reported that they could not make the session. Seventy-six percent of felt that it was very important that they could bring their partner, and 95% of those whose partner attended thought that the class was useful to their birth partner. Most women (93%) stated that they would recommend to a friend that they bring a birth partner with them.

When comparing ACE with other classes, 58% of women and 52% of birth partners planned to attend other classes, with the most popular being hypnobirthing (37% women, 24% birth partners). When comparing ACE and NHS classes only 11 women and 5 partners had attended NHS classes prior to their ACE session, preventing meaningful conclusions from being drawn. However, participants indicated that they received similar information and that both were useful. However, if they had to pick one class the majority of participants stated that they would select the ACE class (W=64%, BPs=60%). After the class there was an increase in the number of women who indicated that they had thought about strategies for coping with labour (91% post vs 78% pre) and knew how to find further answers to questions about labour and birth (89% post vs 64% pre).

#### Feedback from Midwives

Midwives generally felt the session went to plan, liked the COVID-imposed small group sizes and the structure. However, they wanted more interaction and time to deliver the class. These data are presented in supplementary file 5.

### Phase 4: Refinement

#### During the intervention

Changes were made in response to the midwife focus group and the feedback from the interviews with parents. These included adding in more explanation of emergencies, rearranging the seating plan and the way the resources were displayed.

#### Post-intervention refinement

We made changes where there were consistent reports from midwives, parents or both that a change should be made. These are displayed in table 2. Overall, 38 changes were recommended: 22 by parents, 5 by midwives, and 11 by both. We have incorporated 36 of these within the training manual. We were unable to address two consistently requested changes: 1) the introduction of a postnatal session, because this was outside the remit of this project, and 2) improvement to the ethnic and socio-economic diversity of the videos, as we were limited by those who were willing to be filmed. We did not have sufficient funding to develop further videos.

## Discussion

In this paper we present the co-design and feasibility work of an optimised two-hour antenatal education session for labour and birth. The ACE birth journeys ‘river’ concept was developed by the co-design groups and delivered by local clinical staff to participants. Through implementation in an NHS setting and evaluation with both participants and providers we systematically identified key issues in the delivery of the session both concurrently and after completion of the classes. We used a collaborative, systematic decision-making approach to addressing issues that were consistently reported by women, midwives or both leading to refinements, to the ACE manual, and the development of a class that can be delivered within the constraints of NHS capacity.

In general women/pregnant people reported enjoying the session, found it informative and useful in helping them to prepare for birth. However, it is notable that partners described being less engaged with the class. Nonetheless, both women/pregnant people and their partners who provided feedback found specific content of the session useful. Women/pregnant people reported increased preparedness, having thought more about coping strategies for labour, and knew how to find further information following the session compared with before attendance.

In this study we have explored potential impacts of the ACE intervention on women/pregnant person’s preparedness and informedness. This contributes to the data on antenatal education interventions, which are heterogenous. They include studies on general antenatal education,^20 21 22^ promotion of self-efficacy^23^ or mindfulness, ^24^ with some focussing on specific topics or outcomes such as preventing postnatal depression,^25^ coping with fear^26^ and breastfeeding.^27 28^ Many of these interventions have been tested in the context of a clinical trial and their length varies from a few hours to multiple sessions over several weeks. These interventions have not been tested within the NHS. Furthermore, given current NHS resource constraints they are unlikely to be feasible to deliver in the context of current care.

There are some key differences between our approach and that of other studies to evaluating the impacts of the ACE intervention in terms of plausible effects of antenatal education. Many studies into antenatal education focus on clinical outcomes, for example, rates of epidural and mode of birth.^29^ We would question whether it is the role of antenatal education to alter mode of birth, and challenge whether value should be placed on reducing or increasing epidural rates – women/pregnant people should be able to select their personal preference.

Our focus was to give information to support preparedness and informed decision making. We believe it may be more beneficial for evaluations of antenatal education to examine outcomes related to what antenatal education should aim to achieve, for example, feelings of preparedness for birth, knowledge of the process of labour and birth, birth expectations and whether their birth has met them, and a sense of empowerment. Elements of existing systematic reviews do suggest that antenatal education can positively impact the labour and birth journey by reducing false labour admissions, which can be stressful for the mother, reduce anxiety and increase partner involvement.^30^ A more recent review^31^ has suggested that antenatal education can impact maternal stress and improve self-efficacy.^31^ A review focusing on childbirth self-efficacy alone suggested that antenatal education promotes women’s self-belief and is effective in achieving a positive birth experience.^23^ Our study did not specifically measure these outcomes, as our aim was to develop and refine the intervention, however it is plausible that this co-designed class, covering general birth preparation within the context of a 2-hour session, could contribute to improving experience of birth for women in resource-constrained environments such as the NHS, and should be the focus of future research.

Of particular importance in our findings was the feedback from birth partners. While they found the information useful, they reported that the class did not meet their needs. This finding is in keeping with existing literature suggesting that birth partners feel outnumbered, excluded, anxious and uncertain and require more targeted birth preparation.^32^ A recent large qualitative study has highlighted the importance of antenatal education in meeting the needs of birth partners,^33^ however there is less literature on exactly how the existing classes could be modified, or new classes designed, to meet the needs of birth partners. While we gathered data from a small number of birth partners during the co-design stage and after evaluation to inform the changes made to the ACE intervention, a limited number of partners took part in them, meaning that it is unclear whether their views were representative. Further research is needed, and should specifically target birth partners, to explore in greater detail their needs to further refine the ACE class.

### Strengths and limitations

An important strength of the ACE intervention is that it was delivered within the allotted two hours, by NHS staff and women/pregnant people felt that the intervention was acceptable and useful. This may make it more feasible to roll out across other NHS Trusts. A further strength is that where possible, we iteratively addressed issues throughout the implementation period, to improve the experience for participants, and we sought feedback from multiple sources to enable us to refine the class. Furthermore, this intervention is underpinned by the experiences of women and pregnant people who have recently given birth.

However, we acknowledge that focus groups undertaken lacked ethnic and socio-economic diversity. We attempted to diversify the sample attending the class by contacting all women greater than 24 weeks of pregnancy booked at the trust on up to two occasions to increase engagement. Despite these efforts we did not achieve representation of women/birthing people that was consistent with our local population. There is evidence of barriers to attendance at antenatal education in women from underserved groups,^34^ therefore further efforts to identify ways to better engage these groups are needed. This may be achieved through public-patient involvement activities to identify barriers and how to address them. Increasing accessibility by providing sessions in local settings may increase attendance and provision of classes in other languages could address potential barriers. A weakness identified by attendees was that the class focussed solely on the labour and birth element of antenatal education, this was due to an existing class in our Trust that specifically addresses infant feeding. Finally, COVID-19 restrictions limited the location of the class and the spacing of attendees within it. A physically larger venue was required which meant classes were held centrally. This may have reduced the opportunity for interaction and relationship forming between participants as they attended the class to suit them, rather than the one held in their locality.

## Conclusion

We have co-designed a structured antenatal education session about labour and birth that provides women/pregnant people with the information they want and need to prepare for birth, within the constraints of available NHS resource. The next step is to ensure that all parents have access to these classes, to support their journey into birth and beyond. To achieve this, national antenatal education guidelines are urgently needed to ensure equitable access to antenatal education.

## Data Availability

All data produced in the present study are available upon reasonable request with appropriate ethical approvals in place.

## Acknowledgements

Our thanks go to all of the staff, women and their partners who supported the development, attended and provided feedback on the session.

## Contribution to authorship

AM conceived the study, obtained funding, designed and carried out study, analysed findings and wrote the first draft of the report. MT, KB, KR, SB, MS, RP, MLa, ADa conceived the study, obtained funding, designed and carried out the study and edited the final draft. MLy designed and carried out the study and edited the final draft. GC, MG analysed the results and edited the final draft. ADe, LW, NM, AC, KB, CD, EH, TRW, EL carried out the study and edited the final draft. CB, AF conceived the study, obtained funding, designed the study and edited the final draft.

