## Supplementary file 1 for "Co-design and refinement of an optimised antenatal education session to better inform women and prepare them for labour and birth"

### Slide 1
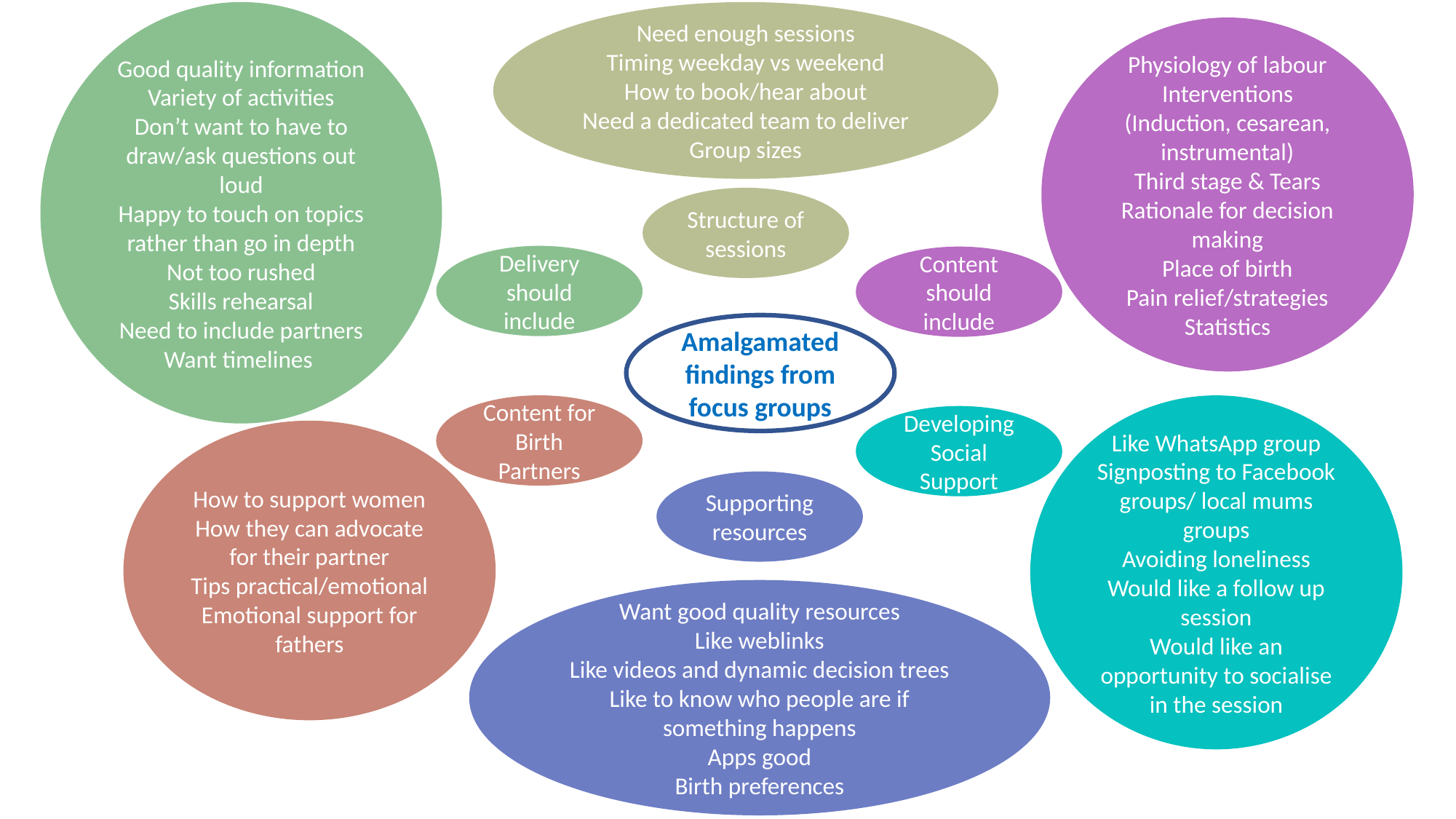

Good quality information
Variety of activities
Don’t want to have to draw/ask questions out loud
Happy to touch on topics rather than go in depth
Not too rushed
Skills rehearsal
Need to include partners
Want timelines
Need enough sessions
Timing weekday vs weekend
How to book/hear about
Need a dedicated team to deliver
Group sizes
Physiology of labour
Interventions (Induction, cesarean, instrumental)
Third stage & Tears
Rationale for decision making
Place of birth
Pain relief/strategies
Statistics
Structure of sessions
Delivery should include
Content should include
Amalgamated findings from focus groups
Content for Birth Partners
Like WhatsApp group
Signposting to Facebook groups/ local mums groups
Avoiding loneliness
Would like a follow up session
Would like an opportunity to socialise in the session
Developing Social Support
How to support women
How they can advocate for their partner
Tips practical/emotional
Emotional support for fathers
Supporting resources
Want good quality resources
Like weblinks
Like videos and dynamic decision trees
Like to know who people are if something happens
Apps good
Birth preferences
