## Supplementary file 2 for "Co-design and refinement of an optimised antenatal education session to better inform women and prepare them for labour and birth"

### BIRTH JOURNEYS

#### Antenatal Education Midwife Manual

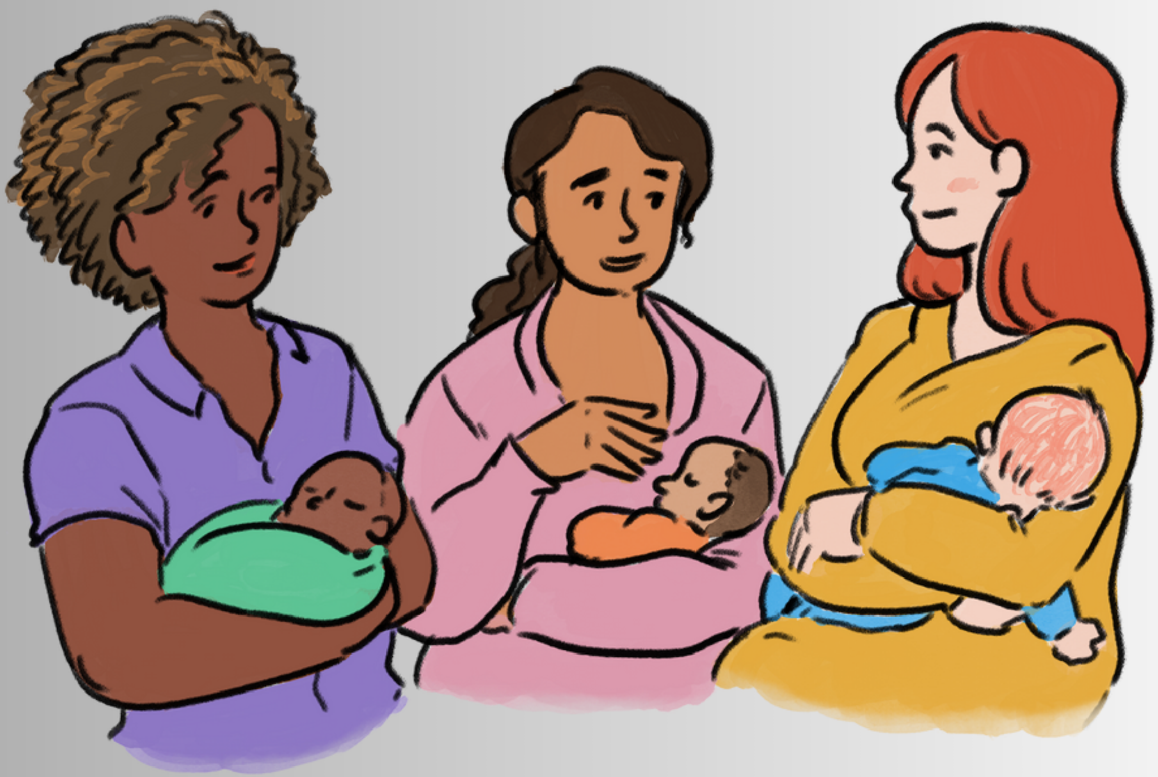

ACE

Antenatal Care Education

WRITTEN BY

The ACE Study Team

University of Bristol
The Health Foundation
NHS North Bristol NHS Trust

ACE
Antenatal Care Education

BIRTH JOURNEYS

### Table of Contents

| | |
|---|---|
| Guide to manual | 01 |
| Resources required to deliver ACE Class | 02 |
| Background to the ACE Antenatal Education Programme | 03 |
| Important lessons for class delivery from our pilot study | 04 |
| Practical lessons for the structure of the classes | 06 |
| Supporting Decision Making | 06 |
| **Section 1:** Introducing the ACE programme and the River Concept | 07 |
| **Section 2:** Birth journeys | 09 |
| **Section 3:** Coping with labour and birth | 21 |
| **Section 4:** Social support for the days and weeks following labour and birth | 27 |
| **Section 5:** Birth preferences | 32 |
| **Section 6:** Answering questions | 34 |
| **Section 7:** Social support opportunity | 34 |
| ***Resources and Appendix*** | 35 |
| ***Glossary*** | 39 |

### Guide to this Manual

This manual supports the implementation of the ‘Birth Journeys’ Antenatal Education Session. It has been co-designed by women and maternity staff based in North Bristol NHS Trust.

The session outlined in this manual is designed to be delivered within a two-hour time frame and focuses on labour and birth.

It is designed to be delivered alongside complementary postnatal/breastfeeding sessions and a session delivered by physiotherapists focusing on keeping healthy during pregnancy, including pelvic floor exercises and coping strategies in labour.

This manual begins with a background to the programme so you can appreciate how the materials were created.

**There are Seven sections within the session:**

**SECTION 1: Introducing the Programme and the River Journey Concept**
**SECTION 2: Birth Journeys**
**SECTION 3: Coping With Labour and Birth**
**SECTION 4: Support Following Birth**
**SECTION 5: Birth Preferences**
**SECTION 6: Answering Questions**
**SECTION 7: Social Opportunity**

We provide a list of resources required for each course component and a suggested timeline for each section.

Throughout the session, we suggest providing some key localised information, including how to find out more about local tours/virtual tours and local birth statistics.

*This manual does not give you a script to use, but it does provide the key points to cover in each section. These key points are based on what women and staff consider important and are also linked to current evidence from peer-reviewed literature and local and national audits.*

### The Resources Required to Implement the ACE Class

This session is designed as a two-hour session about labour and birth alone. It does not address anything other than the most immediate postnatal period. The course resources are available from.

Feedback from midwives suggested that it would be an easier session to deliver in pairs, to facilitate both setting up the session and increasing interaction and to ensure that all of the content is delivered at pace – as two hours is tight.

Whilst it is resource-lite, it will require some investment in the time to deliver sessions and some basic resources.

**Most of the resources can be reused; they include:**

- *Portable projector/laptop and speakers.*
- *Printed laminated birth journeys poster and laminated, blue-tacked birth events/strategies stickers.*
- *You may require a board to stick this onto, depending on your poster/local facilities.*
- *Printed posters for labour progress, cervical dilatation and the 'what's in the bag' exercise if you choose to use them.*
- *Handouts, if you choose to use them.*
- *Flipchart paper / Marker pens if you choose to use them.*
- *Post-it notes if you choose to use them.*
- *Lanyards with pre-printed labels with clinical roles.*
- *Kiwi/forceps/ventouse if available.*

### Background to the ACE Antenatal Education Programme

The ACE (Antenatal Care Education) programme is a Health Foundation-funded quality improvement project focusing on the labour and birth element of antenatal education. It has been developed by drawing upon the diverse experiences of over 50 women/birthing parents and birth partners, as well as clinicians, to identify key strengths and weaknesses in the current antenatal education provided in North Bristol Trust.

The strengths and weaknesses identified in focus groups were used as a foundation by groups of women/birthing parents, staff and the project team, who developed this co-designed programme.

Below are some key findings from the focus group summary:

- Women/birthing parents frequently seek information from the NHS and private/ non-profit organisations (e.g. NCT, Daisy Birthing, Hypnobirthing) about labour and birth. They also report seeking non-NHS education to develop social connections with other women/birthing parents who will be having a baby at the same time.

- A mismatch between the information that women received in these classes and their labour and birth experiences was identified, including:

    - Differences in the pace of labour from what they expected- both considerably shorter or longer than they had understood it to be;

    - The disproportionate focus on spontaneous vaginal birth in the NHS (often referred to as a 'normal' birth) and a lack of information or 'glossing over' of commonly experienced interventions;

    - A lack of understanding of the rates/ frequency of interventions used during labour and birth (e.g. induction, caesarean and assisted birth);

    - A lack of knowledge and misunderstanding of the rationale for some interventions;

    - Limited knowledge about what would happen in the third stage of labour.

- Women/birthing parents demonstrated good knowledge and use of coping strategies that they had learned about and had used during their births and made recommendations about key topics to cover.

- Women/birthing parents reported that they wanted the class lead to cover set topics, with an opportunity to ask questions either privately or in the group. They did not want to generate their own topics and did not wish to be asked to talk or participate in a group where they did not know the other attendees.

- Women/birthing parents reported that they did not find it useful to get information from group activities where there was the opportunity for incorrect information to be shared between attendees. They wanted all group activities to be designed in such a way that they only access correct information to avoid confusion.

- Women/birthing parents appreciated learning about labour and birth through different media to bring to life topics they wished to learn about, for example, the use of interactive materials, such as models, sorting tasks, videos and real-life experiences.

- Women/birthing parents reported wanting to have a sense of the timeline of the process of birth and labour and understand key variations within that timeline.

- Women/birthing parents reported that language is very important. This relates to 'normal/natural birth' and 'elective' and 'emergency' caesarean birth. The terms were viewed as emotive and impacted their emotional response to the situations that they found themselves in. Consider language when discussing birth. Try to use 'birth' instead of 'delivery' and planned and unplanned when discussing caesarean birth.

- Women/birthing parents reported that the NHS class context rarely enabled them to build important social support networks for the days and weeks following the birth. Assistance in developing social connections with other mothers/ birthing people was very important to them.

- Women/birthing parents understood the time constraints of the classes and recommended signposting to detailed, verified resources where time did not allow in-depth discussion.

- A focus on partner support was important, both in relation to being a source of strength to the women in labour and also to helping the birth partners cope with their birth experiences in the days and weeks immediately following birth.

### Important lessons for class delivery from our pilot study

- Women/birthing parents highlighted the importance of using empowering language throughout the class.
- The need to take every opportunity to involve birth partners/ highlight the important role birth partners play.
- It is important to integrate the river concept, see page 7, throughout your session, so it may be useful to practice this with props before delivering a class in person.
- Women/birthing parents appreciate an opportunity to discuss coping strategies and, in particular non-pharmacological pain relief strategies throughout the river journey. When discussing this topic, please highlight that 'coping strategies' can be considered non-pharmacological pain relief and can often continue to be used alongside pharmacological pain relief and in situations where there is some concern about you and your baby.
- We suggest that you read this manual, watch the training video and plan how you will deliver the sessions before your first session. Ideally, you would attend a session delivered by another midwife or deliver a session in a pair whilst you build confidence. This will be dictated by your local resources.
- Women/birthing parents appreciated seeing a kiwi (which is what we had available), but we received feedback suggesting that showing forceps would also have been useful.
- It is useful to highlight that the information presented during the class will be in line with local and national guidelines.

### Practical Lessons for the Structure of the Classes

- If possible, offer classes on different days/times of day to facilitate attendance.
- Please try to set the class up so participants can see any screen and each other. This facilitates discussion, so around a table or in a semi-circle.
- Please highlight to the group that this section covers labour and birth and explain the local provision for other elements of antenatal education.
- Please include at least two interactive activities which get participants on their feet (several are suggested throughout the manual).
- Please include a short break (ideally 2x 5min breaks).
- Please have videos ready to play, and ensure you are familiar with the equipment if you choose to use them.

### Supporting Decision Making

Supporting women/birthing parents in their decision-making is a key aspect of pregnancy and intrapartum care, women/birthing parents who have more of a sense of control through active decision-making have a better experience. It is, therefore, important to discuss during the class at some point how decisions/options can be approached.

One method commonly used is BRAIN:

- **B**enefits - What are the benefits of making this decision?
- **R**isks - What are the risks associated with this decision?
- **A**lternatives - Are there any alternatives?
- **I**ntuition - How do I feel? What does my 'gut' tell me?
- **N**othing - What if I decide to do nothing/wait and see? What happens next?

We would encourage you to share this tool at some point during the class when it feels appropriate.

### SECTION 1: Introducing the Programme and the River Poster

**Objectives of this section**

- **To welcome and make participants feel comfortable.**
- **To give an overview of this session in the context of the other sessions, clarifying which topics are and are not covered and which will be addressed in other sessions.**
- **To introduce the idea of the river journey.**
- **To introduce the key idea that the mother and baby are healthy at the end of the journey as the driving force for women/birthing parents and clinical staff.**

**Resources**

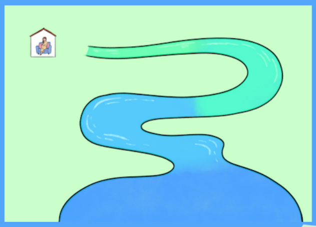

##### 1.1 Welcome the group

Welcome the group to the session, introduce yourself and cover venue housekeeping.

If the group is of an appropriate size (e.g. 5-10), you may wish to ask them to introduce themselves and share some information, e.g. whether the sex of the baby is known or where they are having the baby.

##### 1.2 Explain what will be covered in the session and how it sits in the context of the overall antenatal education programme

Familiarise yourself with the planned sessions, e.g. when the Physiotherapy and breastfeeding sessions are. Explain the structure of the locally available sessions and how this fits in.

Describe that this session will give facts and information about labour and birth, what to expect, how to cope with it and signpost to information they may find useful.

##### 1.3 Outline of the session

- Birth Journeys:
    1. What a spontaneous vaginal birth is like
    2. Assisted birth
    3. Caesarean birth
    4. Induction of labour
- Coping with labour and birth
- Social Support following birth
- Preparing for birth
- Answering questions
- Social opportunity (add in wherever suits)

##### 1.4 Questions:

It is important that you decide how you would like to deal with questions. Some participants will be happy just to ask freely aloud, but others would prefer an alternative way to ask questions. Women have reported in our focus groups that they don't like having to shout questions out to a group. Explain that you will come back to them all at the end. You may consider post-it notes and invite people to write down their questions and put them up in a specific place during the short breaks, such as on a flip chart or into a pot. You may suggest that people can stay after the class to ask you their questions. Whatever you decide to do, ensure that you have the correct resources available and explain clearly to women at the start of the session how you will address questions.

##### 1.5 Introduce the River concept

- Use the river to introduce the idea of labour and birth as a journey.
- You start at home before you go into labour or when you go into labour.
- Your journey will end with you having a baby surrounded by family, friends and support from the community if you need to access it.
- There is a current that gives you momentum through your journey.
- There are many different currents that you could encounter in the river, but all are going in the same direction.
- There are choices/decisions that you can make along the way.
- At all times, the goals of having a healthy mother/birthing person and baby are of primary importance to everyone involved.

##### 1.6 Introduce Birth Preferences / Circle of Importance

Introduce them to the Birth Preference tool. See details in Section 5.

### SECTION 2: Birth Journeys

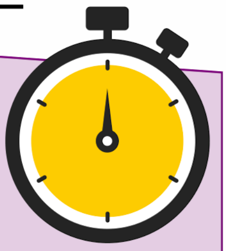

#### Objectives of this section

To introduce the different types of birth.
To demonstrate the likelihood of each type of birth.
To demonstrate the reasons for different types of intervention.
To demonstrate clear, factual information about the stages of labour, what happens physiologically and what a woman/ birthing parent will feel in each stage.
To inform women using the river about the points at which different interventions can occur.

**POSTER TIP: We struggled to get posters to stick onto the wall. You could overcome this by pinning it onto a notice board. We bought a A0 sized canvas and an easel to stand the poster on to make this easier.**

#### Resources

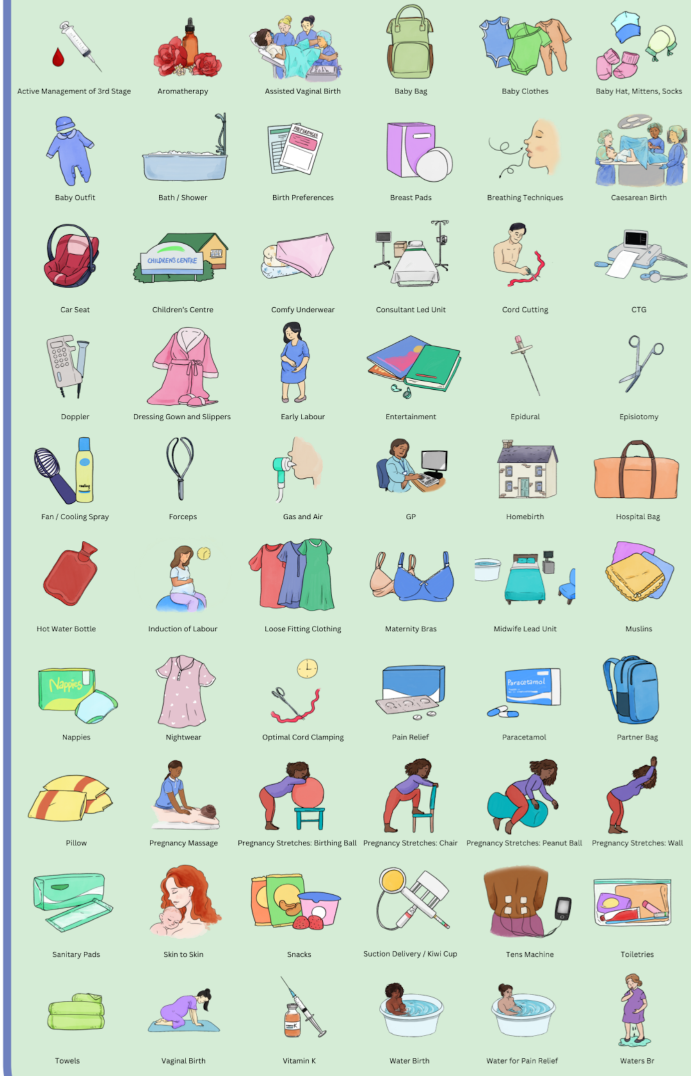


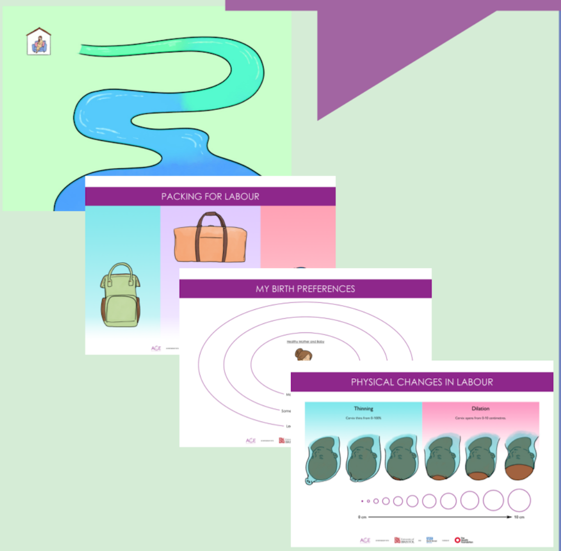


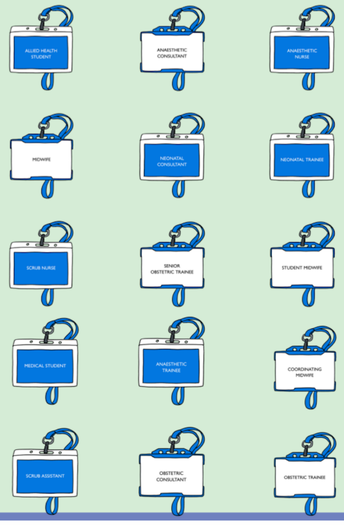

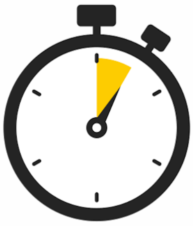

**Introduce the different places for birth:**

Explain the different locations for birth (can use stick-on pictures for home/midwife-led unit/consultant-led unit).

Explain a bit about what can drive the location.

- Choice
- Past history
- Current pregnancy factors

Explain that there is sometimes a need to change the place of birth during labour/birth due to additional information/needing something different. This could be between 35 and 45% for women/birthing parents having their first baby but is much lower (about 10%) in a subsequent pregnancy.

Explain when the place of birth discussion takes place/who with.

Signpost to how women/birthing parents can access more information/tours/virtual tours about the trust's local options.

**Introduce the topic of types of birth:**

Preparation Task: Demonstrate what proportion of women/birthing parents in the room will have a straightforward birth and the likelihood of interventions.

- Before the start of the session, check your local maternity statistics for mode of birth.
- Roughly mentally divide the participants in the room.
- At the start of each of the sections below, communicate those proportions by asking groups of people in the room to stand up to indicate the proportion of women/ birthing parents who have different types of birth.

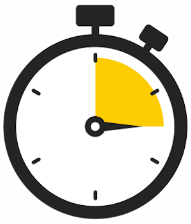

#### TOPIC 1: Spontaneous Vaginal Birth

**Suggested Interactive Activity:**

Ask the group to split into two/three groups. Get them to write on a sheet what to put in a hospital bag for one minute. Ask the groups to give feedback and 'pack' the hospital bag resource poster as they feedback -adding in any missed items.

These are Mother/birthing parent - Baby - Birthpartner bags for you to use as you see fit and a range of stickers with items to pack in each bag.

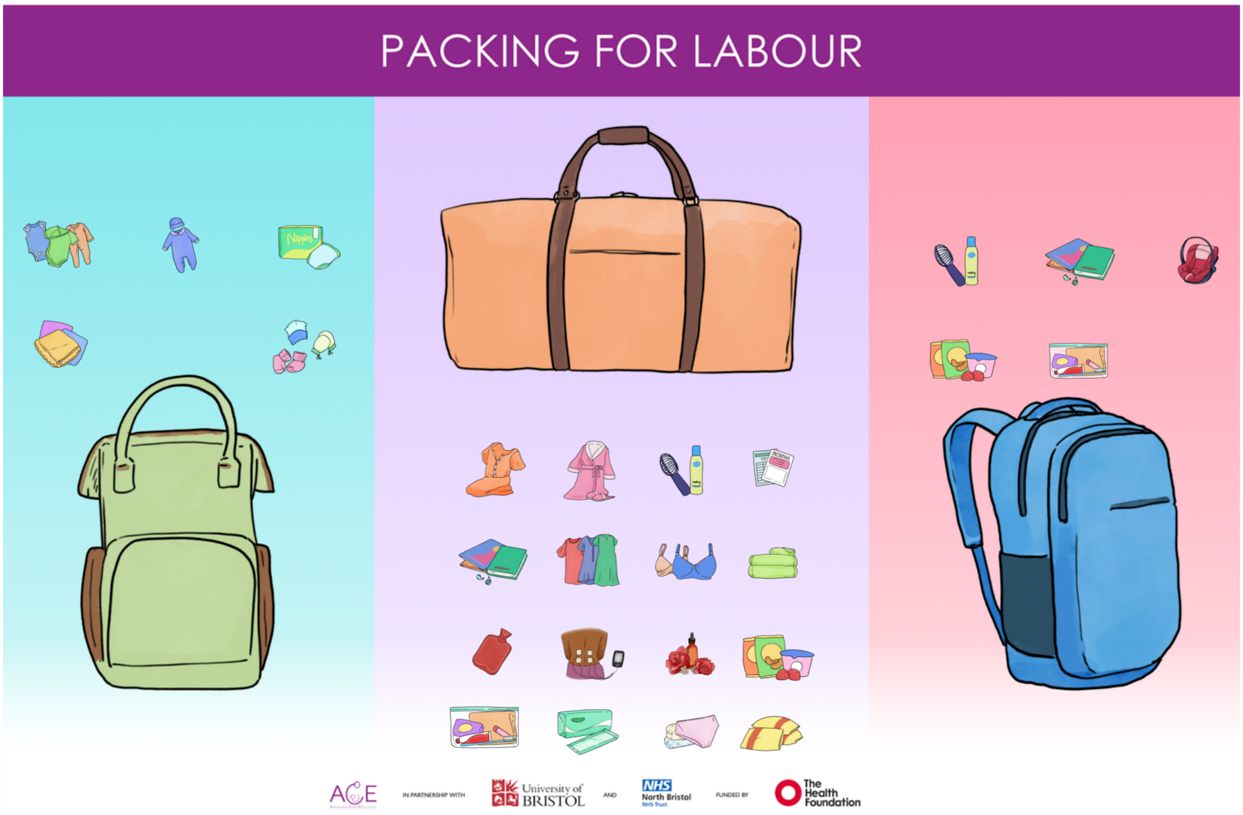


***Resources:***

- *Circle of Importance/ Birth Preferences.*

**Preparing for Birth:**

- Opportunity to prepare practically (packing bag, getting tools for coping ready (discussed later)).
- Consider options and choices.
- Introduce the circle of importance activity, which can be used throughout the session.

**Activity: See the detailed Instructions in section 5**

- To support birth preparation, a circle of importance tool allows note-making of where their preferences lie during the session.

- Healthy mother/ birthing parent and baby are in the centre – to depict that this is important to all.
- Describe how the things most important to them are at the centre, and then position them outwards from there as we move through the session/ consider afterwards at home.

**The Stages of Labour:**

- Use the pictures to stick on and explain where events could happen. Sometimes you need several copies of the same laminated sticker.
- Run through the stages of labour using the river to help depict the journey and the timeline.
- The river is divided into four colours.
    - 1st
    - 2nd
    - 3rd
    - Post Natal

**Latent Phase: Right at the start of the journey at home**

- Describe the latent phase.
- Importance of calling in and coming in to be assessed.
- Discuss how the waters may break and need to contact a midwife/ planned birth setting.

**First Stage: At home and then in the hospital**

- Labours are all different and of different durations.
- Describe the first stage and relate it to the team's resources.
- Assessment is important, and when to call in.
- Intrapartum fetal monitoring – dependent on the level of risk of pregnancy and on the setting; please ensure discussion about Intermittent auscultation and electronic Fetal Monitoring and the difference between them.

**Second Stage**

- Passive and active phases.
- Monitoring (Intermittent Auscultation and Electronic Fetal Monitoring).
- Length of time.
- Vaginal examinations to assess progress.
- Perineal Protection for Prevention of Tears (OASI Project).
- Sometimes, episiotomy is needed.

**Third Stage**

- Optimised cord clamping and cutting.
- Delivery of the placenta.
- Active/physiological 3rd stage with a focus on a recommendation for the active third stage to reduce blood loss and the need for blood transfusion (see RCOG PPH Guideline).
- Assessment for tears – discuss that tears are common.
- Repair of tear: analgesia (local/epidural/spinal).
- Remind them that some of these things are choices they can make, and therefore they may consider sticking them to their birth preferences.

**The Baby**

- Some babies need some help after birth, especially with breathing.
- Even babies born at term can still end up in the neonatal unit for some support.
- If an instrumental delivery/caesarean birth is not planned, a baby doctor will be present at birth.
- Baby will be offered Vitamin K orally/ IM -explain the pros and cons.
- Please mention that some babies can be born very unwell and need a log of support from baby doctors. It is more likely that they are born early or have problems with how the baby is put together.  But cannot predict all those needing input or having bad oucomes.

**Emergencies:**

Explain that sometimes there is an urgent need to intervene during or immediately after birth for the sake of the mother and child. If you experience this during or after a vaginal birth, it may be that your midwife will press the emergency button, and the room fills up with people.

***Suggested Activity:*** *Give everyone  in the class a role to show who will be in the room if there's an emergency (can also save the exercise for later during the caesarean birth part).*

Explain briefly the commonest obstetric emergencies without trying to make them alarming, for example:

- It may be that your baby isn't getting enough oxygen, sometimes when this happens, the baby's heartbeat is low; this will mean that the doctor will want to help you to have your baby quickly unless we can use other measures to help increase your baby's heart rate quickly. This may mean that the doctor will offer a caesarean or assisted vaginal birth -whichever is quicker, which will happen very quickly.

- It may be that once your baby's head is born, its shoulders don't want to come out easily; this is called shoulder dystocia. This means that lots of people will come into the room, and we will need to put your legs into a funny position to deliver your baby; this can be uncomfortable. Still, we need to move quickly so your baby gets enough oxygen.

- Sometimes women bleed after birth, and this can mean that we need to give you extra drugs to make your womb squeeze and stop bleeding. It is likely also to mean that you need to have an internal examination and have your tummy rubbed, and have a catheter inserted. Lots of people will be in the room, and sometimes you need to go to theatre.

- Sometimes when babies are born, they may need help with breathing. This can be the case with spontaneous birth but is more likely if there is an emergency. We can sometimes need to get babies into a good position and open their airways. Sometimes, because they have been breathing fluid, we need to help them to clear their lungs too. If we need some support from the baby doctors, we will call them to come and assist, and several may arrive at once.

- For any of these emergencies, if you are not in a hospital, it is likely that you will need to go there either for you or your baby.

**Case Vignettes/ Videos:**

You can show or share stories from women who have had two different kinds of vaginal birth (see appendix).

**This is what the river may look like at the end of this section.**

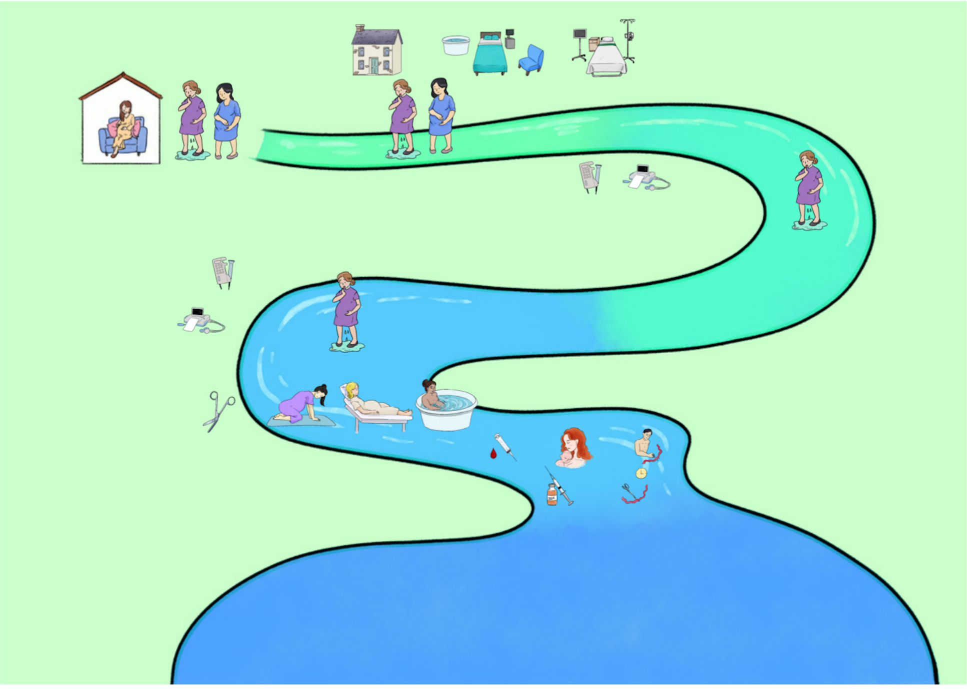

#### TOPIC 2: Assisted Vaginal Birth

***Resources required:***

- *Video/audio/written vignette of instrumental birth experience.*
- *Blue-tacked instrument, all delivery stickers.*
- *Kiwi/ventouse/forceps if possible.*

**Likelihood of Instrumental Birth**

**Activity:**

Proportions task: Ask the approximate proportion of the room to stand up 'This number of people from this room will have an assisted vaginal birth (USE CURRENT FIGURE from local maternity dashboard).

**Reasons for Instrumental Birth**

- Planned: due to medical history and the need not to push.
- Urgent: Baby distressed.
- Length of labour: very long labour, meaning mum/ birthing parent is exhausted or increases the risk of bleeding after birth.

**The process of an Assisted Vaginal Birth**

- Acknowledge that assisted vaginal birth deliveries are a known source of concern among women/birthing parents.
- Indicate on the river when an assisted vaginal birth usually occurs using stick-on pictures.
- Mention the types of assisted vaginal birth: forceps and ventouse (suction cup/vacuum).
- Key point: The choice of the instrument lies with the doctor. The decision of what instrument is complex, taking in factors to do with mothers/birthing parents, baby and analgesia. If you decide you do not want forceps, this may mean that assisted vaginal birth is not an option for you. Instead, a caesarean birth will be offered. It may well be that the quickest and safest way to have your baby is with an assisted vaginal birth.
- Discuss pain relief used: local/epidural/spinal.
- Discuss the use of episiotomy – episiotomy is not routine but at the discretion of the operator to create space for the birth of a baby and also to try to protect your back passage from a tear.

**The Process of Assisted Vaginal Birth (Continued)**

- Discuss who is usually in the room during birth – dependent on location (in room 1-2 doctors, a baby doctor (paediatrician), at least two midwives, a maternity assistant in theatre, there is also at least one anaesthetist and one anaesthetic assistant at a minimum).
- Discuss the place of assisted vaginal birth: labour ward and theatre.
- If assisted vaginal birth is not successful – quick change to caesarean birth.
- What parts of an assisted vaginal birth are the same or similar to a spontaneous vaginal birth?
    - Vaginal birth.
    - Mum/birthing parent is doing most of the work (i.e. pushing).
    - Usually, a Partner can cut the cord if they wish.
    - Skin to skin either immediately or once the baby doctor is happy if needed.

Emphasise that there are still choices that can be made in terms of the above and the team do their very best to facilitate this.  There are some elements they may wish to add to their birth preparation tool in relation to this.

Present vignette/video of assisted vaginal delivery experience (see appendix).

**This is what the river may look like.**

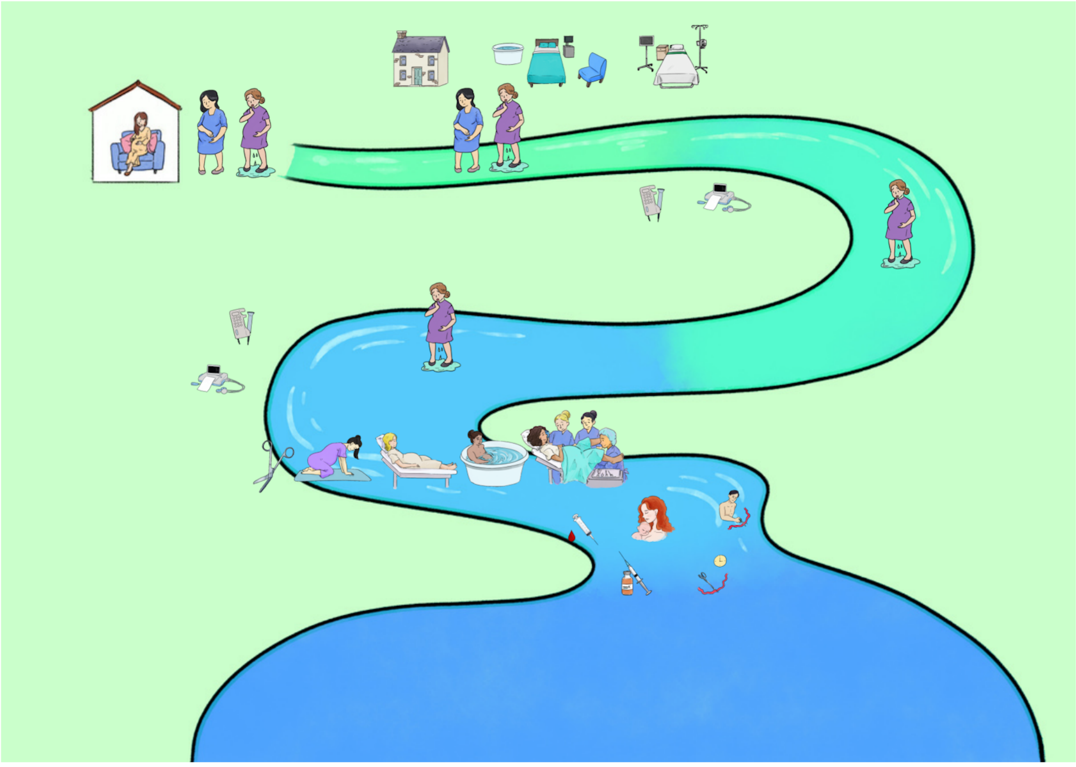



ACE
Antenatal Care Education

### TOPIC 3: Caesarean Birth

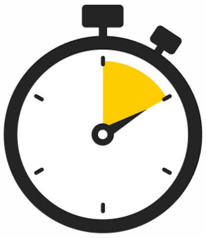

***Resources required:***

- *Blue-tacked caesarean stickers.*
- *Lanyards for 'who is in the room' activity.*
- *Video/audio/written case vignettes.*

**Likelihood of Caesarean Birth**

**Activity:** Proportions task: Ask the approximate proportion of the room to stand up. This number of people from this room will have a.......planned caesarean, unplanned caesarean. Get the proportions from the local maternity dashboard.

**Reasons for Caesarian Birth**

- For what reasons is a planned caesarean offered? They are referred to as 'elective' caesareans but note that this language can mislead because many/most are planned as a response to present risk factors. Medical issues, previous surgery/Caesarian, maternal request, planned delivery for a complication, e.g. placenta/growth.
- For what reasons is an unplanned caesarean offered: baby distress, maternal issues, e.g. bleeding, slow progress in labour, unable to get into labour, maternal request.

**The Process of a Planned Caesarean Birth**

- Using the stickers, indicate where on the river a planned caesarean occurs.
- Describe the process of a planned caesarean to include prior appointments to discuss and consent, attending on the day, spinal block, the procedure, time in hospital, and catheter.

**The Process of an Unplanned Caesarean Birth**

- Clarify language around an unplanned caesarean- some might not be very urgent, and others can be extremely urgent.
- Indicate using the stick-on pictures where on the river an unplanned caesarean can occur in the Latent, 1st, 2nd stage, or pre-labour.
- Describe the process of an unplanned caesarean, including spinal block/epidural top-up and the operation itself.

**What parts of having a Caesarean are similar to a Spontaneous Vaginal Birth**

- Delayed cord clamping.
- Partners can usually trim the cord if they want to.
- Dropping the drape to see the baby/the birth partner can announce the sex.
- Early skin-to-skin with you/partner if wanted.
- Choice of music in theatre.

**Activity: Who is in the room during a Caesarean Birth?**
**(If not done earlier)**

- Ask for 10-12 volunteers.
- For each volunteer, indicate who they are and what their role is in the operating theatre using a lanyard.
- Indicate that this can be surprising and that this number of people is standard care for caesarean birth.

**This is what the river may look like at the end of this section.**

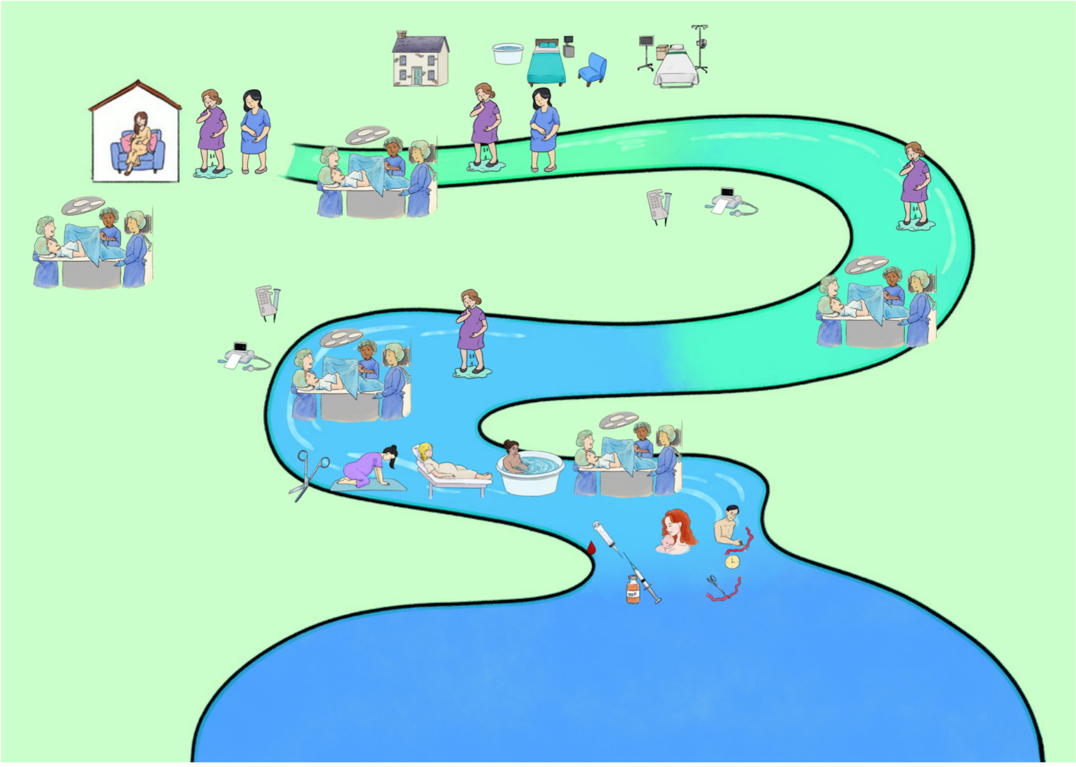

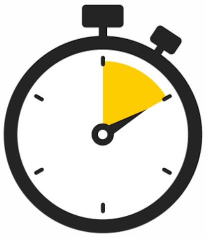

### TOPIC 4: Induction of Labour

***Resources required:***

- *Video/audio/written case vignette of induction stories.*
- *Stick on induction pictures.*

**Likelihood and Reasons for Induction**

**Activity: Proportions task**

Ask the approximate proportion of the room to stand up. ‘This number of people from this room will have an induction. Get the proportions from the local maternity dashboard.

For what reasons is induction offered?

- Planned (past history, medical issues, e.g. diabetes, age, request).
- Emerging (growth, gestational diabetes, waters breaking, reduced movements at term, request).

**The Process of Induction**

**Resources required:**

- Demonstrate on the river when an induction might occur using the pictures.
- Indicate that it can be used to start labour or can happen after the waters have broken.
- Outpatient and inpatient, according to local protocols.
- Describe the process of induction relating to the different types of induction.
- Describe the timeline of Induction, including the potential for it to take several days and the reasons for this.
- Indicate the importance of planning parental leave where necessary.

**Induction Impacts**

- Reduces Unplanned Caesarian rates and no proven reduction in spontaneous birth rates (in a randomised trial of induction vs expectant management in low-risk nulliparous women/birthing parents, there is no difference between the two groups).
- Reduced Perinatal Mortality, some increased admission to neonatal units, but no increase in poor outcomes. e.g. babies born needing support with breathing/brain injury.
- It is likely to mean longer in the hospital and the labour ward.
- It is likely to mean you cannot deliver on the midwife-led units (however, in some units, this may be facilitated if Syntocinon is not required).

**This is what the river may look like at the end of this section.**

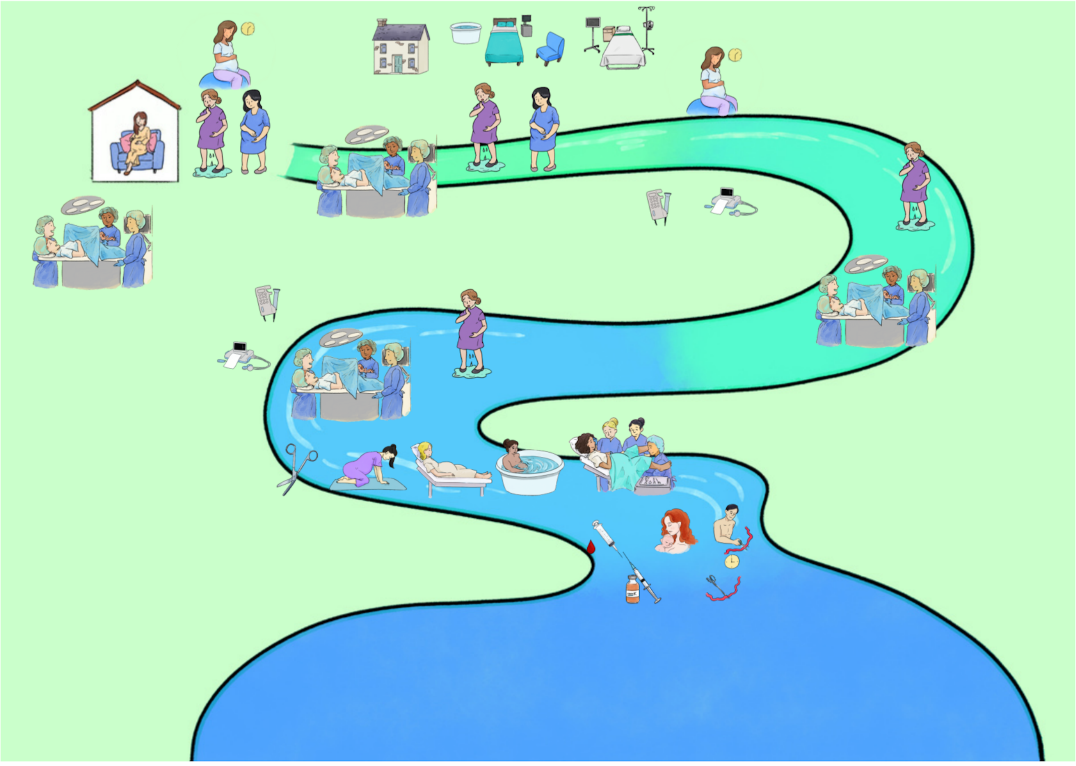

### SECTION 3: Coping with Labour and Birth

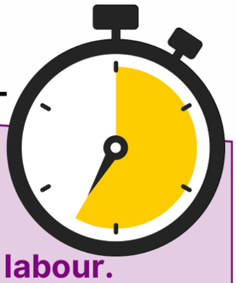

**Objectives of this section**

**To outline the different coping strategies that can be used during labour.**
**To outline the pain relief options available to women/birthing parents.**
**To signpost women/birthing parents to the physiotherapy session.**
**To outline the partner's role in providing support through labour and birth.**

#### Resources

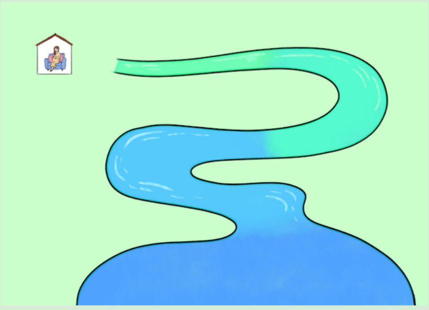

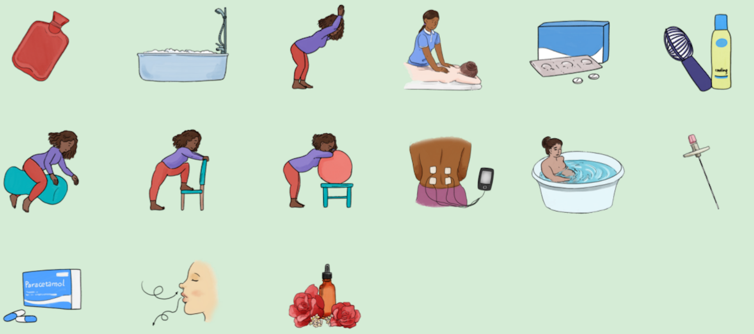

**Introduce the topic:**

- This section prepares women/birthing parents with coping strategies for labour and birth.
- We will use the river to show the different places where each coping mechanism can be used.
- This element will not cover any area in detail –but we will signpost you to further local information.
- Highlight any resources available locally for hire, e.g. tens machines from the birth centre.
- Encourage attendance at other antenatal education sessions if your trust has them (e.g. postnatal/infant feeding/physio).

### TOPIC 1: Preparing for Labour and Birth

**Activity: Building a Latent Phase Toolkit**

Suggest what people may like to prepare to help at home. e.g. hot-water bottle, tens machine, bouncy birthing ball.
Discuss these areas and others and stick them onto the river journey at the relevant points.

**During Labour:**

Discuss the above strategies and add other strategies, including water, movement, positions, breathing, hypnobirthing, relaxation techniques, making the room your own, music to take with you/have ready at home, aromatherapy, massage...

**Suggested Exercise: Breathing Exercises**

Explain that it can be useful to use pre-learned breathing exercises to deal with pain in labour and that these exercises can be used at any time during labour and birth, even if medical interactions are needed.

***You can then explain the technique you teach, which may include:***

- Touching fingertips together at the bottom of the ribcage, breathing in for five and out for five and seeing how fingertips move apart – this trains for deep breathing.
- Sign out slowly, 'Breathe in through your nose and sigh out through your mouth' (in 2,3 out 2,3).
- Focus on the out-breath, and the breath will come in automatically.

**This is what the river may look like at the end of this section.**

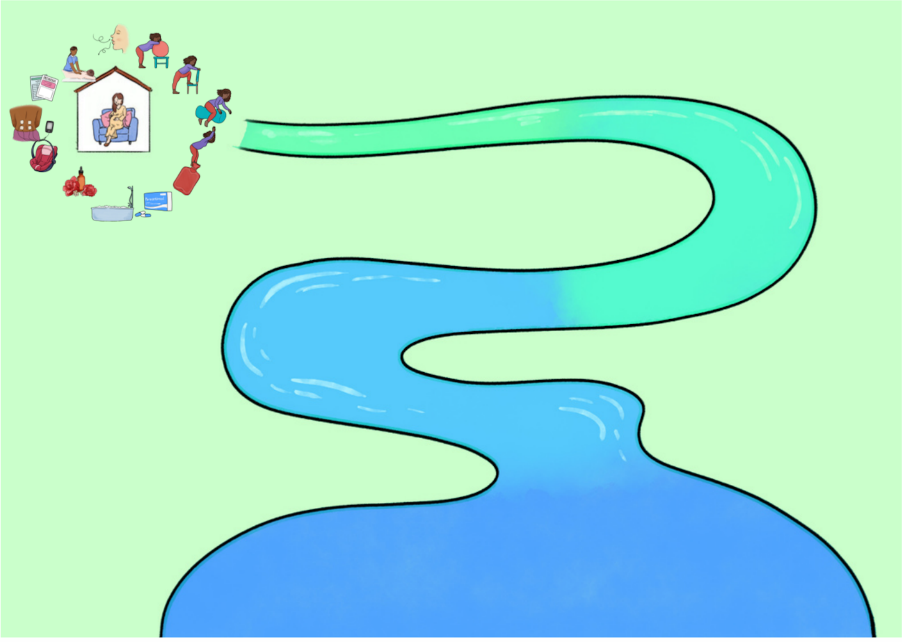



Antenatal Care Education

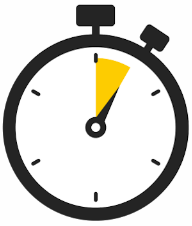

### TOPIC 2: Pain Relief Options

**Activity: Discussion of pain relief options**

**Paracetamol & Dihydrocodeine.**

**Gas and Air** can help reduce pain, doesn't tend to take it all away; breathe it in with contractions, and it works in 15-20 seconds – it also leaves the system quickly, so if you don't like it/make feel funny, the effects will end quickly.

**Pethidine/Morphine** injection (given with anti-sickness) lasts 2-4 hours, can make you drowsy and can make the baby sleepy. We don't often advise it after 7-8cm as it can cross the placenta and make the baby tired at birth. Breastfeeding may be immediately affected if the baby is sleepy when born.

**Epidural (patient controlled)** This is an injection into the back with a small plastic tube left in the back so that it is possible to continue to inject some local anaesthetic to numb the pain by blocking the nerves. The benefit is that for most women/birthing parents, the labour pains are entirely removed.  There are some possible risks, including severe headaches after birth, which may need to be treated with another injection in your back and a rare complication of nerve damage.

**This is what the river may look like at the end of this section.**

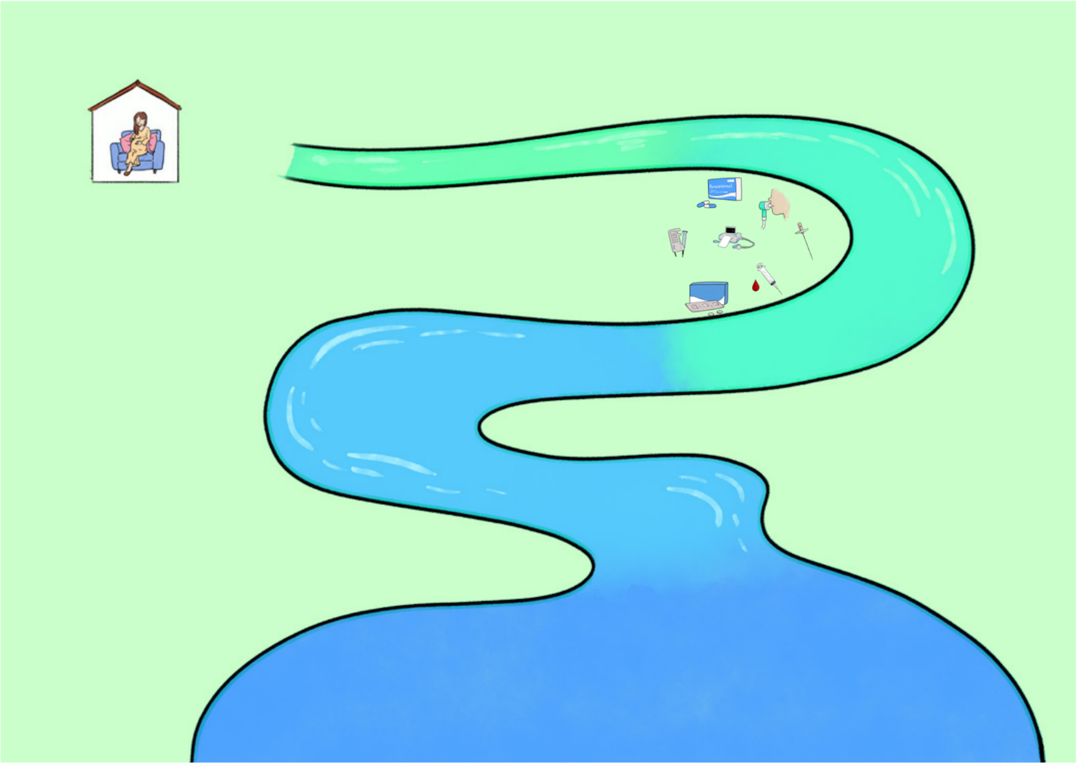

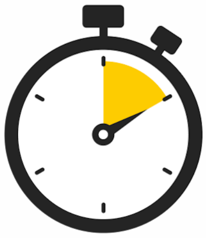

#### TOPIC 3: Non-Pharmacological Coping Strategies

**Please acknowledge that many of these things are not evidence-based and not recommended by NICE guidelines perse.**

**Things to discuss include:**

**Breathing Techniques – Hypnobirthing.**

Many midwives are trained to support women in labour using these techniques.
There may also be local classes for parents in preparation for labour.

**Aromatherapy**

Often midwives are trained to use essential oils.

**Massage**

Some midwives are trained in this to support physiological & psychological well-being for women reducing anxiety and promoting calm, comfort and connection for pregnancy, birth and beyond.

**Birthing Ball**

Supportive during contractions- sway/rock/move in the rhythm of contractions.
Adopt different positions during labour – leaning/hands & knees.

**SESSION TIP: This session could be carried out as suggestions from the floor.**

**Birthing Peanut Balls**

Useful for adopting various positions during labour.
It assists in opening the pelvic outlet & helpful in allowing babies to rotate to a good position during the course of labour.

**TENS Machine**

Transcutaneous Electrical Nerve Stimulation.
Sends small, safe pulses of electrical current via the leads to the pads on the skin. Supportive as pain relief in early stages of labour.
Available to hire as well as purchase.

**Pool**

Discuss water as a coping strategy and where pools are available.

**This is what the river may look like at the end of this section.**

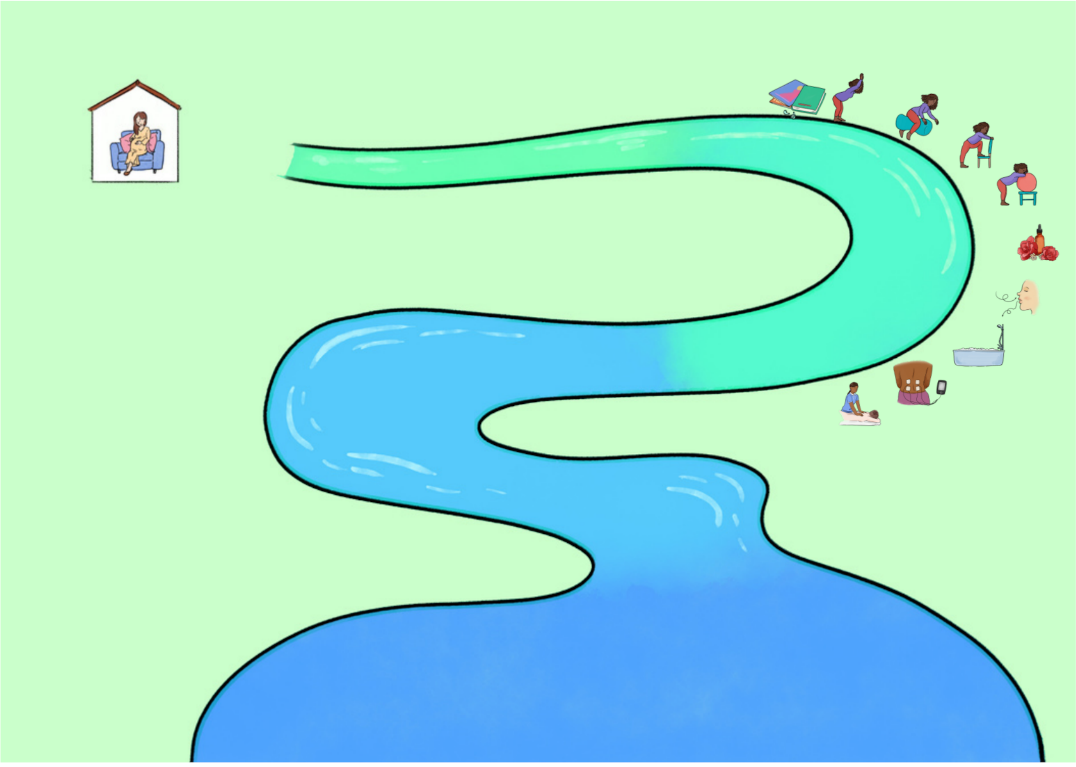



ACE
Antenatal Care Education

### TOPIC 4: Support from Your Partner

**Practical:**

- Preparing snacks and fluids (although be mindful to drink to thirst only and drinking isotonic drinks can be helpful).
- Preparing music.
- Knowing where everything is in the hospital bag, especially the hat and nappy.
- Photographer.

**Physical:**

- Rubbing back/ massage (especially if you have decided on massage techniques).
- Supporting with moving around/position changes (especially if you have practised positions).
- Meeting your partner's needs – some women/ birthing parents like no contact in labour, others really like it – just being supportive of what they want.

**Emotional:**

- Being emotionally present in the room, e.g. not on the phone all the time.
- Support partner in pre-planned wishes but also be supportive of the evolving situation and changes in plans.

**SESSION TIP: This session could be carried out as suggestions from the floor.**

### SECTION 4: Support for the days and weeks following birth

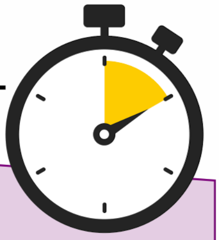

**Objectives of this section**

**To briefly address caring for oneself after birth: physical and emotional.**
**To highlight the appointments and services that are part of standard care.**
**To signpost possible support services following birth if more support is needed.**
**To indicate the benefit of making a social connection.**

#### Resources

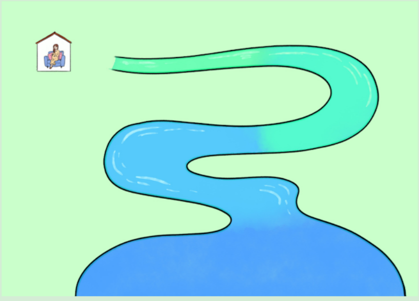

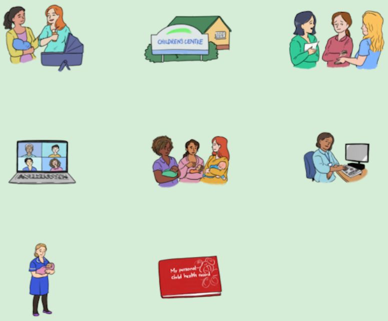

**Introduce the Topic:**

- The focus of this session is not to cover the postnatal period, but several attendees expressed a desire to have some information about the postnatal period.

- It is necessary to offer a further postnatal session as this section is not designed to address the topics in any great detail.

#### TOPIC 1: Caring for yourself After Birth

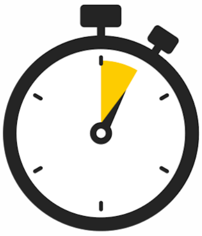

**Days after birth: acknowledge that this is brief information:**

- Healing and pain – taking regular painkillers, being kind to yourself, and not expecting to be back to usual straight away.
- Healing – whether this is vaginal or caesarean birth takes time. Then can explain absorbable sutures and sometimes non-absorbable ones following caesarian.
- Going to the toilet opening bowels (many women/birthing parents are concerned about this – give some top tips including laxatives if stitches, positioning with legs up).
- Going for a wee – explain that you can go in a bath or pour water over the perineum simultaneously to dilute the urine.
- Explain the need to keep any wound clean and dry.
- Emotions: baby blues, post-natal depression spectrum.
- These things are normal – ask the advice of the hospital team/community midwives.

**Discuss ways women/ birthing parents can look after themselves:**

- Perineal care.
- Physical things (not lifting, getting back to exercise slowly, sleeping when the baby sleeps).
- Eating healthily (postnatal – cake and coffee = weight gain).
- Emotional adjustment to life, including relationship adjustment.
- Baby blues (completely normal and last a few days) – emotional/ irritable/ anxious/ restless.

**Activity: proportions task:**

Ask the 10% of the room to stand up 'This number of people from this room will have postnatal depression.'

Postnatal depression (1/10 women/birthing parents ) – loss of interest in baby/ feeling hopeless/ can't stop crying/ not being able to cope / not enjoying things /can't concentrate / memory loss / excessive anxiety.

#### TOPIC 2: Standard Care

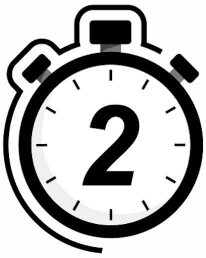

**Describe the Routine:**

Midwife appointments and their different locations.

**Explain How:**

Health visitors take over care and methods of contact.

The baby check with the GP.

#### TOPIC 3: Support Services

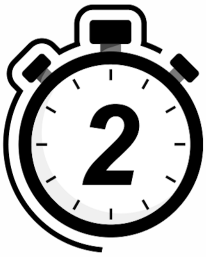

Stick on to the harbour area of the river drawing the following labels:

Health visitors, midwives, GP, local services and children's centres.

**This is what the river may look like at the end of this section.**

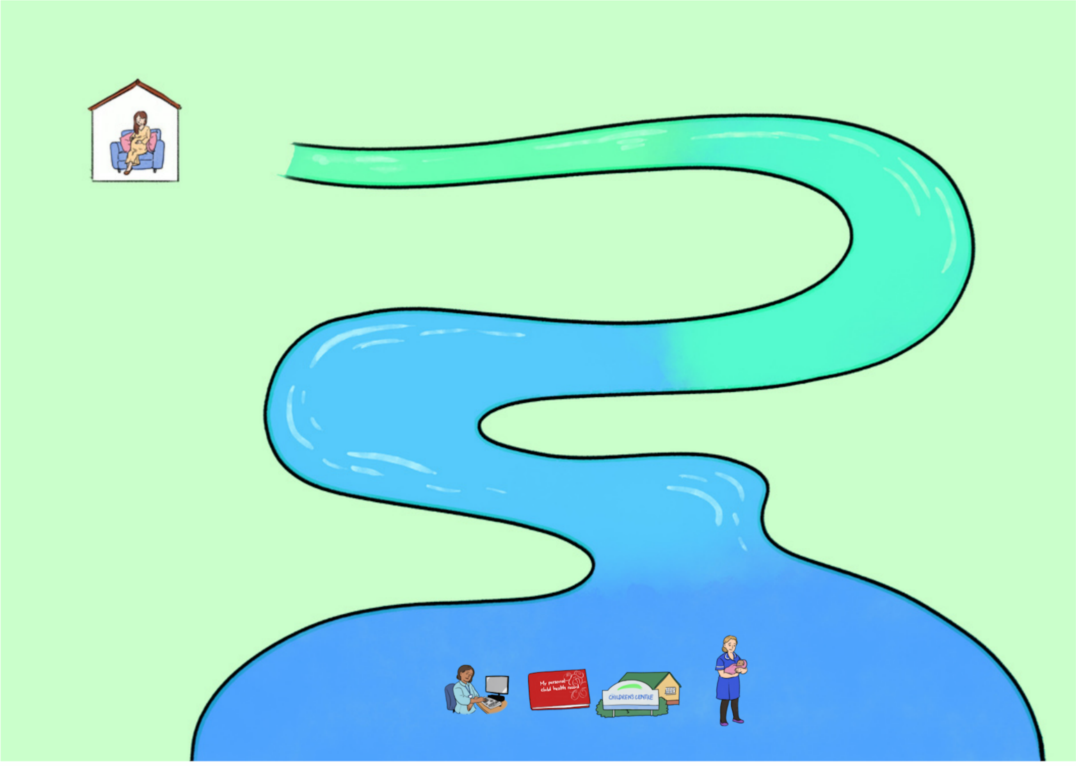

#### TOPIC 4: Social Connections

- Highlight the importance of this and how you hope people from this group may want to stay in touch.
- Avoidance of isolation is important.
- Support from peers around all aspects of being a new parent is important.
- Local breastfeeding support groups on Facebook.
- Local baby/toddler groups.
- Highly encourage engagement in these.

**This is what the river may look like at the end of this section**.

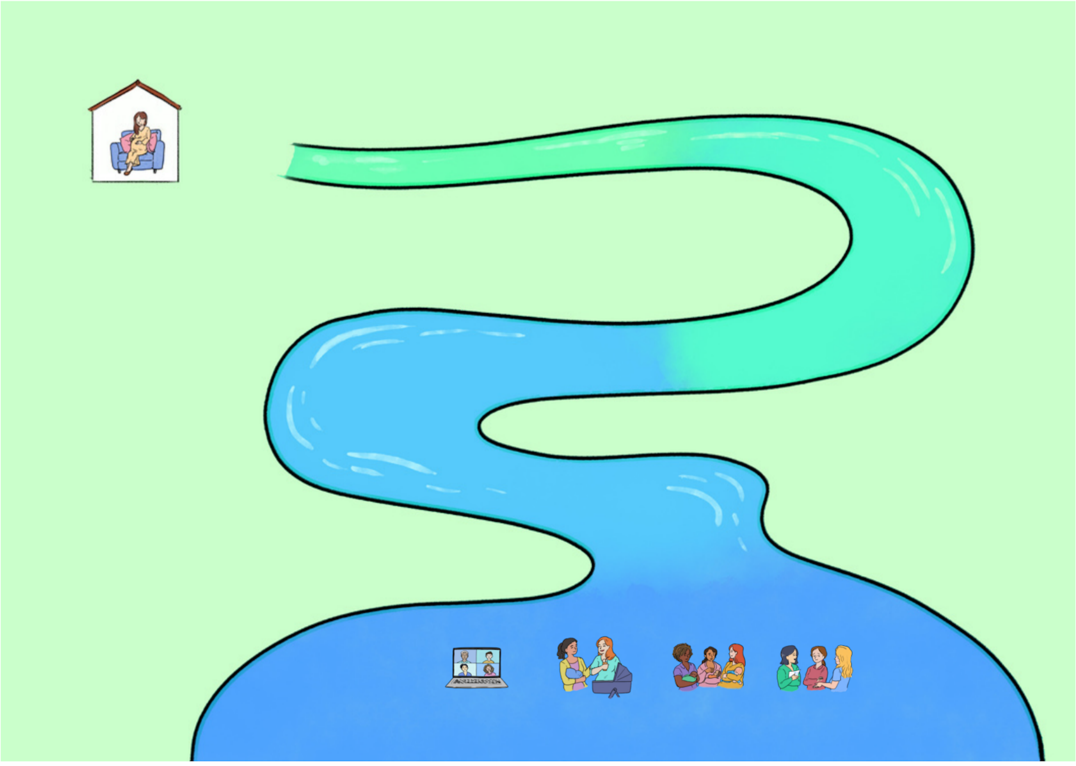

### River Poster

**This is what the river may look like at the end of the whole section with everything on the poster.**

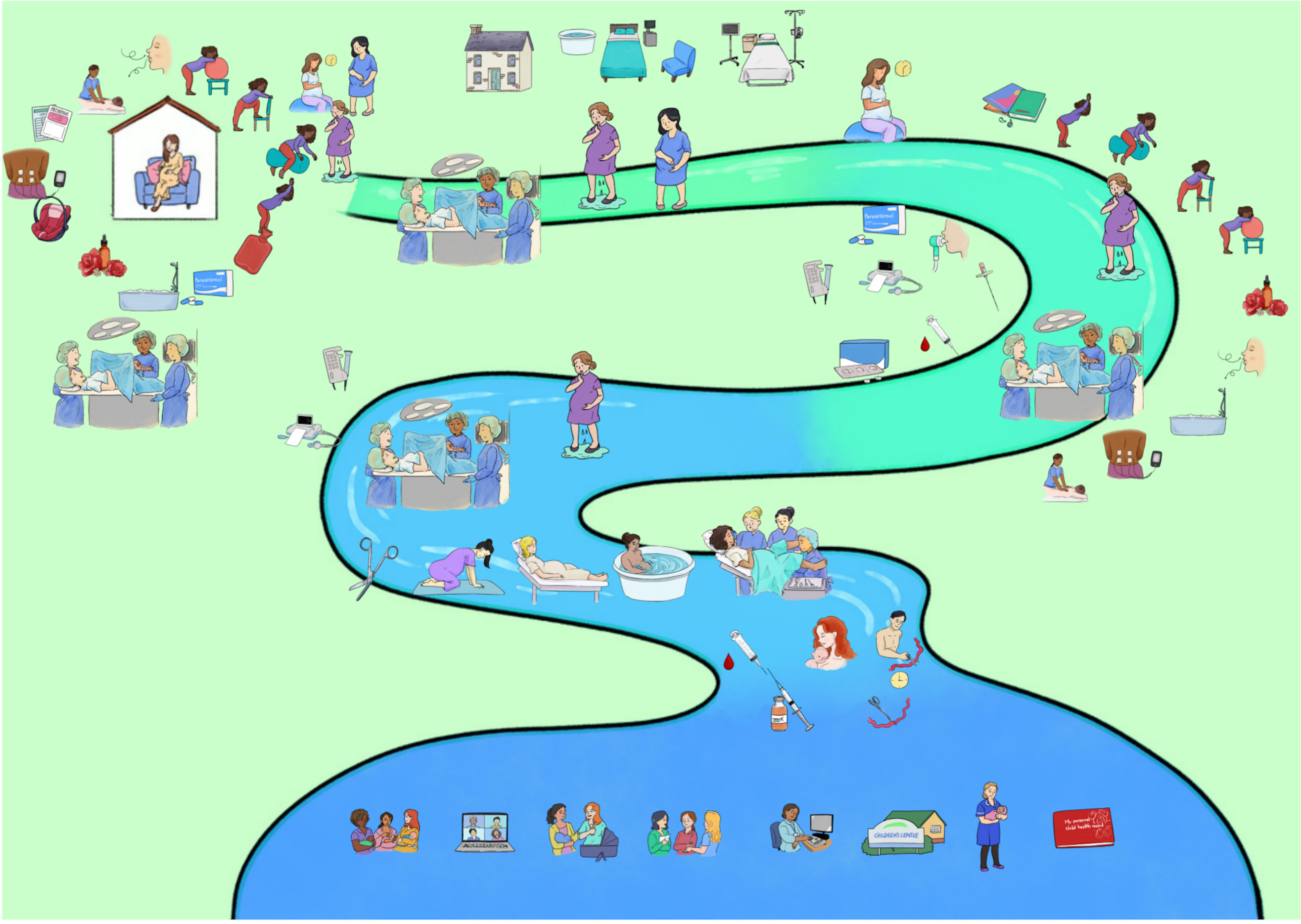

### SECTION 5: Birth preferences

##### Objectives of this section

**To discuss the role of a birth preferences tool.**
**To provide one way of considering options for labour and birth.**

#### Resources

- **Printed sheets depending on your preference as a facilitator.**
- **Locally available resources.**

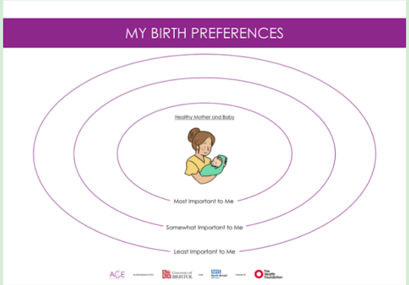


**Things to highlight:**

Discuss the role of birth preferences – promoting consideration of options and how to make choices, but the importance of flexibility.

Discuss the locally available birth preference options for example the online NHS tool, any tool within local pregnancy notes and when the midwifery appointment is in which birth preferences are discussed.

Introduce the idea of considering ‘what is important to you’ when thinking about birth and using these concentric circles as a possible tool to guide thoughts about the various choices around labour and birth. Highlight that at the centre of this is a healthy mother and baby.

**SESSION TIP:**
**At the start of the session, can discuss this to frame the way that parents think about the various options open to them.**

**Towards the end highlight again once lots of the areas have been discussed.**

Make some suggestions as to where to put the elements that have/will be discussed. For example, You may decide that you prefer to use non-pharmacological coping strategies only. Therefore you could put this idea in the centre, or you may decide that you're not very bothered about what pain relief you use during labour as long as you are coping, so this could be categorised in the 'least important to me' circle.

**Opportunity for an activity:**

1. You could divide into small groups to discuss some of the things they may include in their preferences.
2. If you introduce it at the start, you could provide couples with this tool and suggest they jot things down as they go through the class.

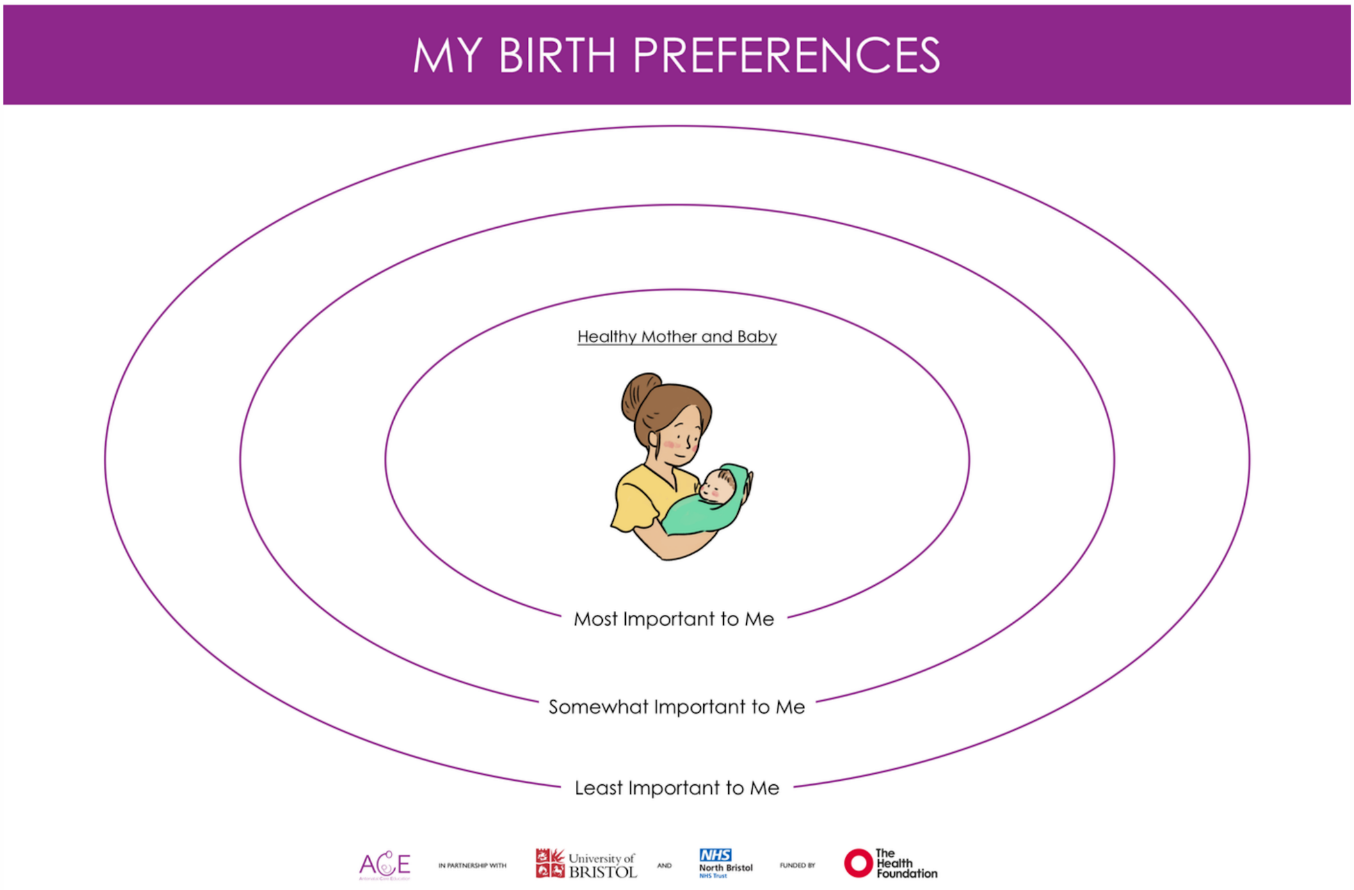

### SECTION 6: Answering Questions

**Objectives of this section**

**To clarify any queries/questions from the group.**

Answer questions before the end – using the system that you explained at the beginning!

### SECTION 7: Social Opportunity

**Objectives of this section**

**To clarify any queries/questions from the group.**

**Pen/pencil and paper**
**Phone**

**Activity: Making connections**

- Collect contact details (with agreement) for a Whatsapp group.
- Find a volunteer to set up a group.
- Encourage participants to go to a local place for coffee if this works within your environment.

**Activity**

Break x2 for 5 Mins.

### Resources

**<u>Appendix 1: Accessing printable images, online training and video case vignettes</u>**

Please for access to the materials.

**<u>Appendix 2: Written Case Vignettes</u>**

**Spontaneous Vaginal Birth 1:**

*I had a long time at home when I was in some pain, I was getting contractions every 10 minutes. It seemed to go on forever, I called up the hospital and came in after a few hours. I wasn't in full blown labour, but they gave me a stretch and sweep, I went home and a few hours later, I was contracting 2/3 times every 10 minutes. I came to the birth suite and it felt like forever. The midwives (I had 2) kept reassuring me that I was making progress and things were going well. They kept listening to my baby's heartbeat and offered me to get in the pool to help with my pain relief. Eventually I was fully dilated. I waited for 1 hour before I started to push and just before I'd been pushing for 2 hours I had my baby who was born straight onto my tummy so I got immediate skin to skin. My partner really didn't want to cut the cord – so the midwife did that – but he did get offered! Oh and the midwife supported the baby's head as he was born and it felt weird, but not uncomfortable. Luckily I just had a small tear which only took a few minutes to fix and apart from some initial discomfort I don't notice now. He was born at 10pm – what a long 48 hours it was for us all – but we went home 11am the next morning and saw the community midwife the next day.*

### APPENDIX

**Spontaneous Vaginal Birth 2:**

*I was well prepared for my long labour and had lots of things ready including a TENS machine and some aromatherapy. After my husband got home from work at 6pm on Friday I started to get some strong period pains, they kept getting closer and closer together and more painful. By 8pm I really couldn't cope with them and when I called the homebirth midwife she said that she would come. She arrived at 8.30 and then assessed me and found that I was 8cm dilated. She immediately called her colleague who arrived about 9.15. Just before she arrived I really wanted to push. It really was very painful but in just a few minutes I had my baby in my arms – I couldn't believe it! I was totally shocked at how quick this was. To be honest, I was a bit sad that I didnt' get to use all of the things I'd planned. I didn't even take my TENS machine out of the box! I had a tear that took a while to repair, the midwife said that my perineum didn't stretch up because it was so quick. But she gave me local anasethetic and gas and air which made it ok.*

**Assisted Vaginal Birth 1:**

*All was great, I was using gas and air and pushing really well the midwife said. But then the baby's heart rate was very low for a few minutes. The midwife told me that she was going to pull the emergency buzzer to get help and that lots of people would come in. I don't really remember but my partner does, it was so dramatic. A doctor introduced themselves I think, and then examined me and told me that I needed an assisted vaginal birth immediately. I said that I didn't want an episiotomy, the doctor explained that they would not do it routinely, but would need to if it was necessary to get the baby out , or avoid a tear of my back passage as the baby's heart rate had now been low for too long. She gave me a quick injection and then put a suction cup on baby's head and the baby was born within a minute or two. The baby was floppy when it was born and went over to the baby doctors. They told me after that she needed some 'inflation breaths' which is apparently very common. Then I got to hold my beautiful girl whilst the doctor fixed the episiotomy. I was uncomfortable for a few days after – but after about 3 days I didn't need painkillers anymore.*

### APPENDIX

**Assisted Vaginal Birth 2:**

*I went to the birth suite when I was in labour. I was contracting once every 5 minutes, it was so painful. The midwife assessed me 4 hours after my first examination. She said I had made some progress, so we kept going. After another 4 hours, I hadn't made any more progress so she thought I would need a hormone drip to help. We moved to the delivery suite and I saw a doctor and the hormone drip started. Things got much more uncomfortable then as I was contracting - my midwife was amazing and helped me with gas and air, positions and pethidine! I got to fully dilated and was so excited. I didn't push for the next hour hour then pushed so hard for two hours. I was exhausted. The doctor came and examined me and explained that the baby was facing sideways. To help her out they needed to turn her around first then guide her out with forceps, although a caesarean was also a possibility. We went to theatre, I had an injection in my back which just made all the pain go away. Then the doctor turned the baby around and used forceps. He gave me a cut and explained why and then my baby was born onto my tummy. My boyfriend cut the cord and the baby never left us. Although he did have some marks on his face for a few hours.*

**Planned Caesarean Birth:**

*We arrived to the maternity unit in the morning, I was a bit hungry and had taken some tablets they had given me. I met the doctors and looked at the consent form again and had a chance to ask any questions. The midwife met me and discussed my options for music, cord cutting, finding out the sex of the baby and whether I wanted the drapes dropped. After that we walked around to the theatre, chatting the whole time, and I met more people in there. They put my music on and I sat on the table and had the injection in my back. I was talking to the anesthetic doctor the whole time, we had a great chat. The operation was odd- and over really quickly! I felt like someone was washing up in my tummy. When the baby was born, I could hear her crying and then they dropped the drapes and my partner took some pictures, then after she had received all of her cord blood, they held her up for me to see! She came straight around to cuddle me. I was really pleased with my whole experience.*

### APPENDIX

**Emergency Caesarean Birth**

*I was in labour, about 3cm dilated I was told. Every time I had a contraction my baby's heart rate dropped. The doctor came in and explained that labour may be tiring the baby. They recommended a caesarean birth. I agreed and they gave me an injection to stop contractions. We were in theatre in minutes. It was a bit odd going from my dim room to a bright theatre, but everyone was lovely and introduced themselves. I had an injection in my back – it was over in moments. Within a few moments my baby was born. I was told she was a bit shocked. She needed some help to breathe at first, but a few minutes later came around to me and I got skin to skin. The doctors talked me through what had happened afterwards and that this didn't mean I needed a caesarean in future.*

**Induction 1**

*I was advised to have an induction of labour because my baby was already predicted to weigh 10 pounds! So at 39 weeks I went to hospital and had the pessary in my vagina for 24 hours. They couldn't break my waters after this, so after waiting a while to see the doctor, they offered to put a balloon through my cervix to gently stretch it. After about 20 hours with the balloon in place, they took me up to labour ward. It was a struggle but they broke my waters and offered me to walk around. I then wasn't in labour so I had a hormone drip. After a couple of hours, I was really in pain, so I decided to have an epidural. Thank goodness I then got some sleep for a few hours. Once I was fully dilated, I waited for 2 hours before pushing and then I had my baby after pushing for almost an hour! It was amazing, she was born and came straight up to me and my husband. I hadn't quite realized how long the induction and labour process was I was in hospital for 50 hours before having my waters broken!*

**Induction 2:**

I decided to opt for an induction because I am 40. I went in and had this thing called a 'propess' a tablet thing on a string that lasts for 24 hours. Within about 3 hours I was in so much pain they pulled out the propess and examined me – I was 5 cm dilated. I was zipped up to labour ward and just half an hour or so later my waters broke. I was 8cm! An hour after that I wanted to push and my son was born less than an hour later. I thought it was all going to be much longer than that!

### Glossary of Pictures

These are the images to be used during the sessions

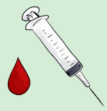
Active Management of 3rd Stage

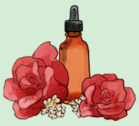
Aromatherapy

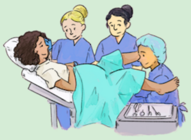
Assisted Vaginal Birth

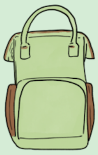
Baby Bag

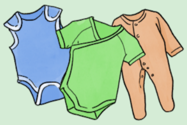
Baby Clothes

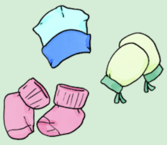
Baby Hat, Mittens, Socks

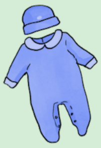
Baby Outfit

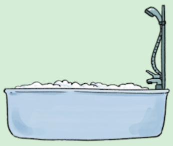
Bath / Shower

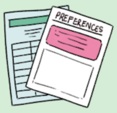

Birth Preferences

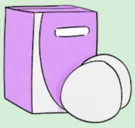
Breast Pads

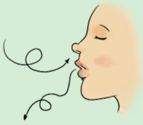
Breathing Techniques

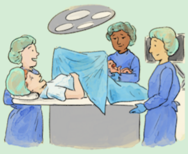
Caesarean Birth

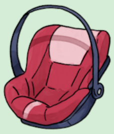
Car Seat

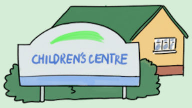

Children's Centre

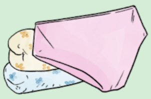
Comfy Underwear

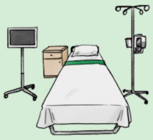
Consultant Led Unit

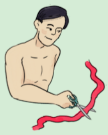
Cord Cutting

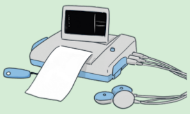
CTG

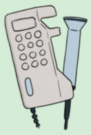
Doppler

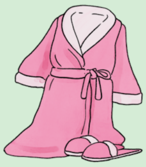
Dressing Gown and Slippers

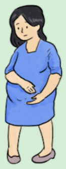
Early Labour

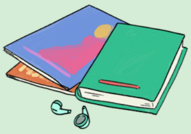
Entertainment

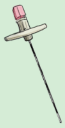
Epidural

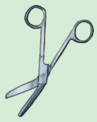
Episiotomy

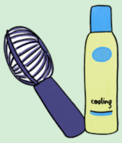

Fan / Cooling Spray

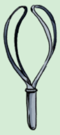
Forceps

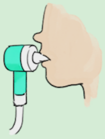
Gas and Air

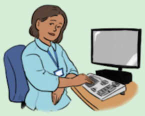
GP

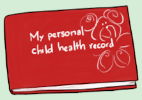

Health Visitor

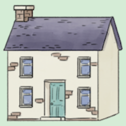
Homebirth

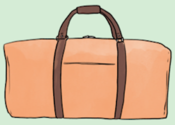
Hospital Bag

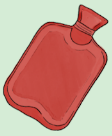
Hot Water Bottle

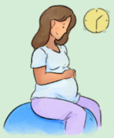
Induction of Labour

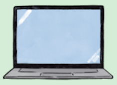
Laptop

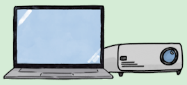
Laptop and Projector

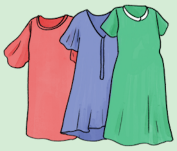
Loose Fitting Clothing

### Glossary of Pictures

These are the images to be used during the sessions

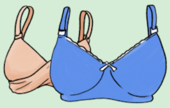
Maternity Bras

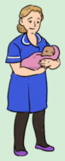
Midwife

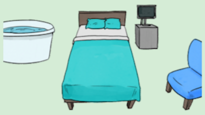
Midwife Lead Unit

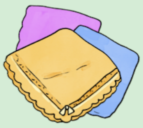
Muslins

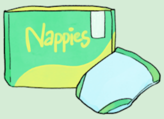

Nappies

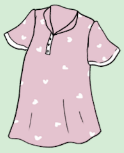
Nightwear

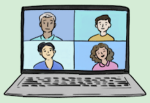
Online Support Groups

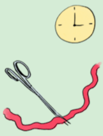
Optimal Cord Clamping

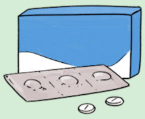
Pain Relief

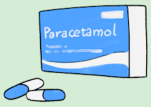

Paracetamol

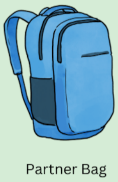
Partner Bag

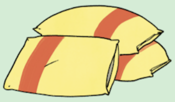
Pillow

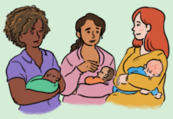
Postnatal Groups

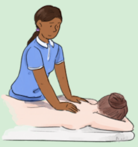
Pregnancy Massage

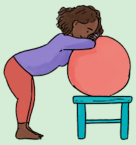
Pregnancy Stretches: Birthing Ball

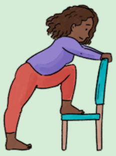
Pregnancy Stretches: Chair

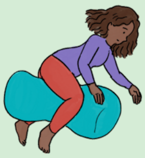
Pregnancy Stretches: Peanut Ball

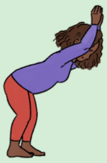
Pregnancy Stretches: Wall

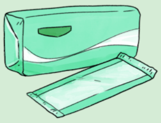
Sanitary Pads

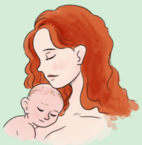
Skin to Skin

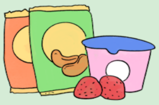
Snacks

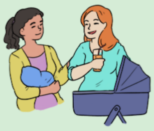
Social Support Network

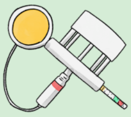
Suction Delivery / Kiwi Cup

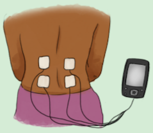
Tens Machine

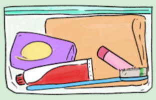
Toiletries

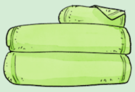
Towels

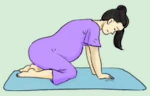
Vaginal Birth

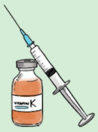
Vitamin K

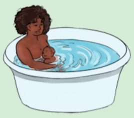
Water Birth

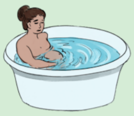
Water for Pain Relief

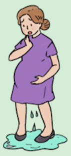
Waters Breaking

ACE
Antenatal Care Education

### Glossary of Lanyards

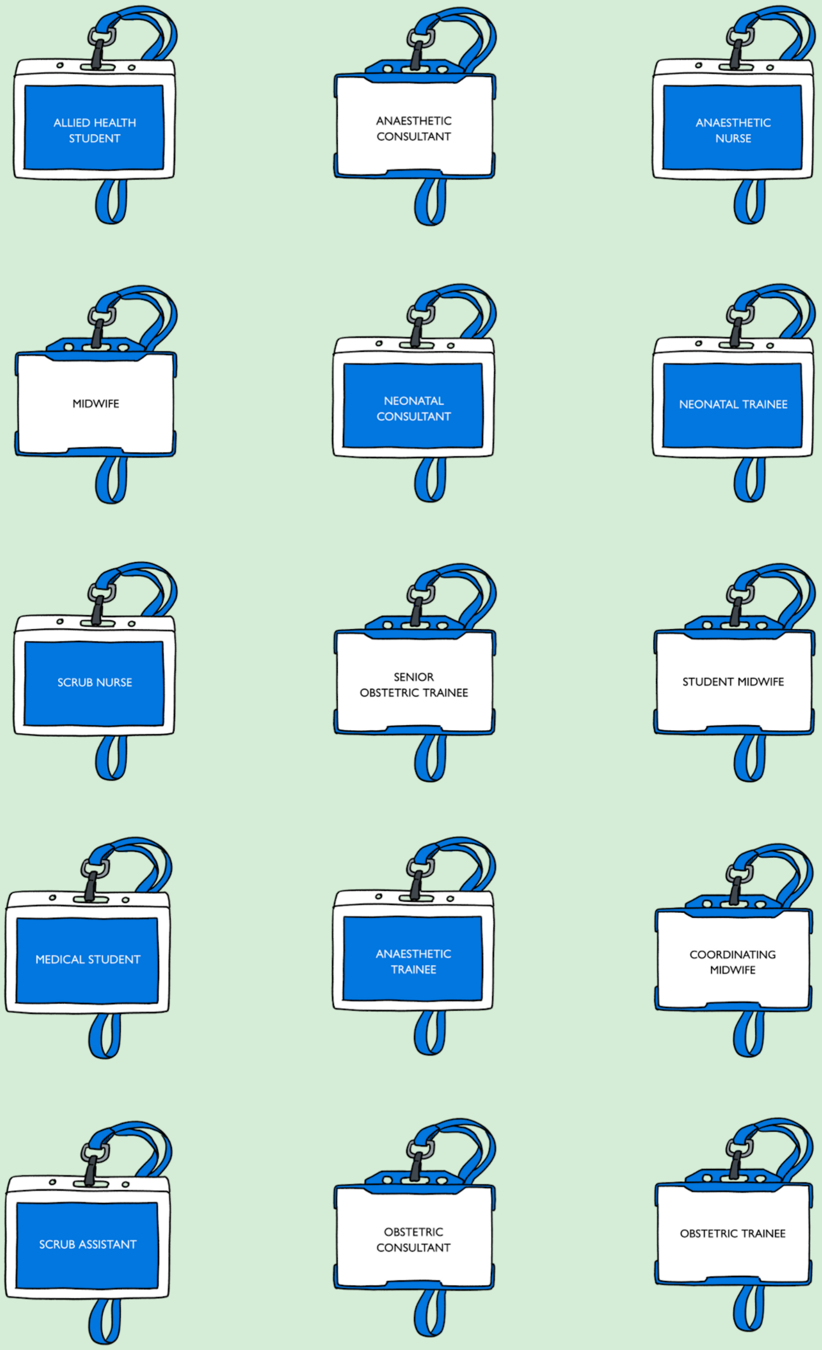

### Posters

You may wish to use these to support the delivery of the class

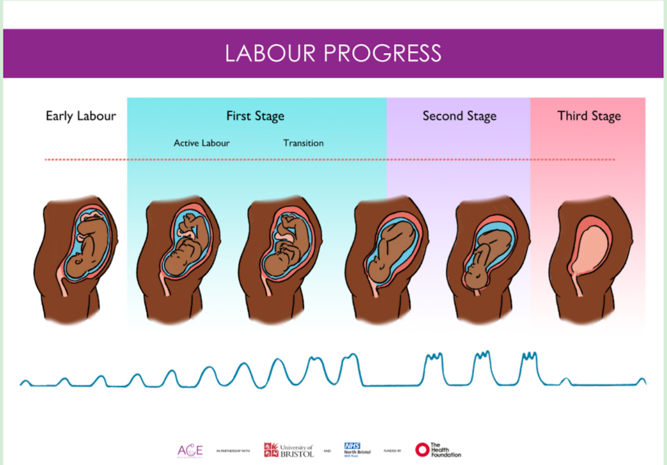


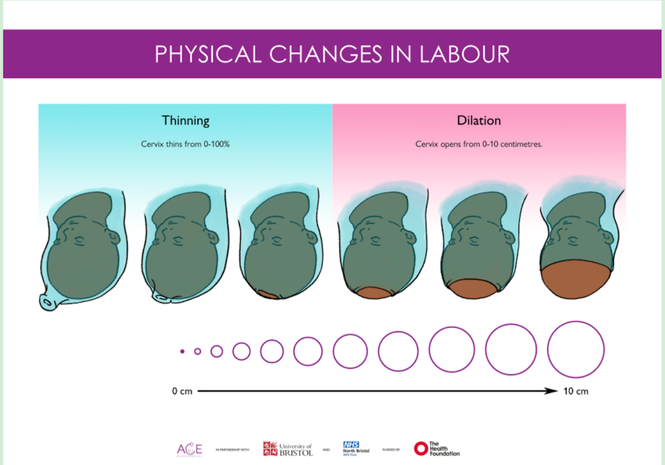


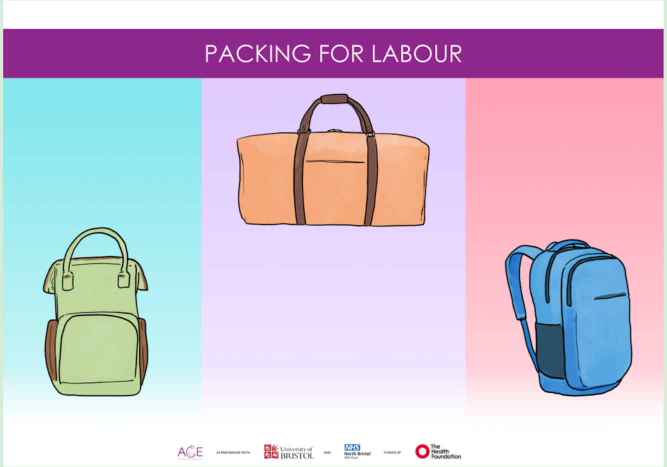


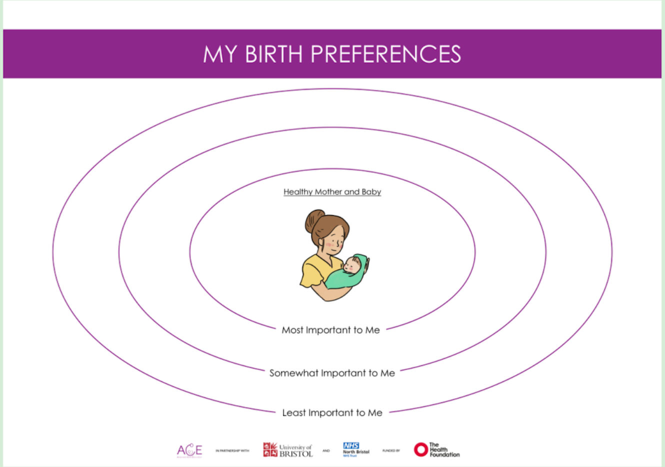


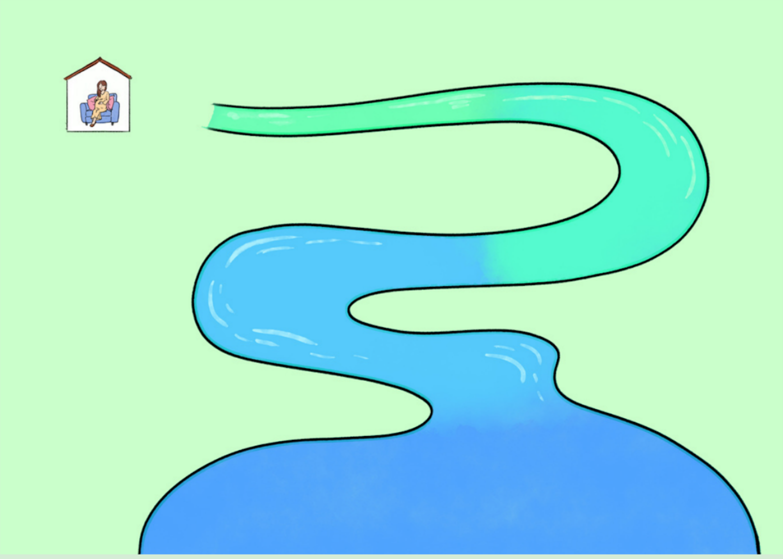

ACE
Antenatal Care Education

ACE

Antenatal Care Education

**Contact:**

**Abi Merriel**

****

University of BRISTOL

The Health Foundation

NHS North Bristol NHS Trust
