## Supplementary file 3 for "Co-design and refinement of an optimised antenatal education session to better inform women and prepare them for labour and birth"

| **Feedback from observed delivery of the 19 ACE sessions** | | | | |
| --- | --- | --- | --- | --- |
| **Topic covered** | **Yes** | **Partially** | **No** | **Not answered** |
| **Section 1: Introducing the ACE programme and the river concept** | 19/19 |  |  |  |
| **Section 2: Birth journeys** |  |  |  |  |
| Topic 1: A straightforward vaginal delivery | 19/19 |  |  |  |
| Topic 2: Assisted delivery with forceps and ventouse | 19/19 |  |  |  |
| Topic 3: Caesarean Section deliveries | 19/19 |  |  |  |
| Topic 4: Induction of labour journey | 17/19 | 2/19 |  |  |
| **Section 3: Coping with labour and birth** |  |  |  |  |
| Topic 1: Coping toolkit for labour and birth | 16/19 | 2/19 |  | 1/19 |
| Topic 2: Pain relief options (pharmacological) | 19/19 |  |  |  |
| Topic 3: Non- pharmacological coping strategies | 18/19 | 1/19 |  |  |
| Topic 4: partner support | 17/19 | 1/19 | 1/19 |  |
| **Section 4:** Social support for the days and weeks following birth |  |  |  |  |
| Topic 1: Caring for yourself after birth | 14/19 |  | 1/19 | 4/19 |
| Topic 2: Standard care | 16/19 | 1/19 |  | 2/19 |
| Topic 3: Support services | 16/19 | 2/19 |  | 1/19 |
| Topic 4: Social Connections | 14/19 | 2/19 |  | 3/19 |
| **Section 5**: Birth preferences | 15/19 | 2/19 |  | 2/19 |
| **Section 6:** Answering questions | 16/19 | 1/19 |  | 2/19 |
| **Section 7:** Social support opportunity | 8/19 |  | 1/19 | 10/19 |
| **Section 8:** Ending the session | 9/19 |  |  | 10/19 |
| **Use of resources** | 15/19 | 2/19 |  | 2/19 |
| **Use of what important to me** | 2/19 | 3/19 | 12/19 | 2/19 |
| **Videos used** | 18/19 |  |  | 1/19 |
| **Participants seemed engaged** | 16/19 | 1/19 |  | 2/19 |
| **Time** | 8 sessions 120 +/- 5 mins  1 session 100 minutes  2 sessions 110-115 mins  3 sessions 40-47 mins over time (same facilitator)  5 session time not recorded | | | |
| **Additional Feedback from observers (general themes):**  Practical issues impacted some sessions (e.g. videos not working easily/late set up)  Timing: some classes over ran significantly  Classes were generally thorough, although occasionally some elements were missed  Questions were asked throughout and groups seemed to interact well | | | | |
