## Supplementary file 4 for "Co-design and refinement of an optimised antenatal education session to better inform women and prepare them for labour and birth"

|  | **Antenatal** | **Postnatal** |
| --- | --- | --- |
| **The value of a class led by an NHS professional in the NHS setting** | ‘It was useful to speak to an actual working midwife, at [hospital name], and understand where there priorities and what their needs are professionally speaking.’ (Mum, 08).  It was good because it was led by a team from [hospital] so it was particularly relevant to us because that’s where we’re hoping to have the baby. So it was really nice to meet the midwives that led the class and get sort of relevant experience and information from them (Mum, 10)  ‘Just them being actual midwives rather than just a facilitator’ (Mum, 010)  ‘Yeah, it just gave me a better idea of what to expect going to that hospital, cost that’s the kind of thing that was missing from the classes before.’ (Mum, 012) | ‘What I really appreciated was that it was midwife led, some of the other stuff I’ve done… but It was midwife led and it was more kind of medical, factual, matter of fact account.’ (Mum, 03)  I’m a physiotherapist, the language I would be more familiar with than some of the people there, so that whole sort of having to cater to  The quality of the information as well, you can go on youTube and google stuff about childbirth but you never know what you’re going to get, the reliability of that information, which as a former health professional I know it’s really important the quality of that information it’s been verified, not just someone making it up on the spot.’ (Dad, 001) |
| **Face to face meeting** | ‘I really appreciated the opportunity to be with somebody live and have that interaction’ (Mum, 001) |  |
| **The river journey** | ‘The journey was very useful, like on the river journey and as you could see the stickers going up you could see how far through the class you were, which I know sounds a bit silly but it was nice to know how you were getting on.’ (Mum, 02).  ‘The only thing I would change about that was it maybe felt like you were saying you’re about to fall into a river, and into a white rapids situation, and if you’re lucky some lifeboat team of medical professionals might be able to come and help you, which is not what she said, it’s not how she put it. But, I think maybe just including something like, if you’ve done your birth plan already, just extend the analogy a bit more, you and your birth partner have designed and built your own raft or boat or whatever else out of the various materials that you’ve found that are important to you, and you’ve got them on this boat, and you might visit these twists and turns in the river on the way, just to feel a bit more empowered’. (Mum, 08)  ‘A big poster about the journey down the river, that was quite nice although it kept falling off the wall. So that was nice, it made it seem a bit more calming, rather than, I’m mean it’s nice when it’s scientific but sometimes it’s nice when it’s a bit sort of softened’. (Mum,010)  I think there was some bits we could have skipped over faster, like the road and the journey (Mum, 11)  ‘I think it would have been better to drop some information like the journey, and focus more on breathing and massage techniques.’ (Mum, 11)  ‘I found the journey helpful because it presented everything in like a chronological order somehow, or in the order you were expecting it to happen (Dad, 11). | ‘There was a river journey, she started using it at the beginning and then it kind of tailed off.’ (Mum, 05)  ‘It kept falling off the wall, I dunno, it was a really nice metaphor of the birth journey, it would have been nice for it to be a bit more interactive.’ (Mum, 24). |
| **Amount of information** | ‘I don’t think it was too much information at all, it was just about right really’ (Mum, 02).  ‘I think it was about right actually, and it looked at it from the right perspective. I can only compare it the class I’ve done before, but that was a bit more technical all about the hormones and stuff and how they worked, and I responded just to this one a lot better. Maybe because it was about what you’d be feeling and what to expect.’ (Mum, 12)  ‘Obviously, otherwise you could talk more, but the class would be four hours.’ (Mum, 13) |  |
| **Prepared for birth** | ‘Some of it I kind of already knew, but I think it was still useful for it to be refreshed, and to confirm that what you’ve read is correct’(Mum, 02)  ‘It was nice to come along even though we’re having a planned caesarean, because it gives me more comfort that should anything happen and the baby comes earlier… it was nice to also know some of these techniques’ (Mum, 02)  ‘As much as I’m trying to feel empowered and informed in terms of the… range of outcomes, I hadn’t realised the number of the medical team so that was really interesting.’ (Who’s in the room task; Mum, 08).  Yes, I think it did actually, and probably more so than the one I attended before. The difference… between me finishing the one I did before and attending the ACE class I’ve been told I probably will need a caesarean, even though a lot of the stuff might not be directly relevant to me, I actually felt it really reassuring to know that if I went into labour before the c-section I know what to expect and how to manage.’ (Mum, 12).  ‘Yes, because having information helps with fear. So you just have less anxiety about things.’ (Mum, 13) | ‘I did [feel prepared] because it was [hospital name] specific, and you mentioned the number of people that would be in, because obviously I ended up having the c-section and I didn’t think I was going to, I remembered from the class all the different people and the number that would be present’ (Mum, 22)  ‘I think it did, it was really informative in terms of the different interventions you could have.’ ‘The detailed stuff about forceps and ventouse’ (Mum, 03)  ‘Well, yeah, cos I had already done the NCT group, and that was more a holistic approach, whereas the ACE one I knew all the things would happen medically and why and when…. I kind of knew what was going to happen next (Mum, 05)  In some ways, obviously the things like discussing the possibility of an induction and sort of what would happen, at least having an awareness of that was helpful was helpful… but if I’m honest, and I don’t know if its how the induction happened and the monitoring issue, I didn’t really feel I knew that electing to have the induction would mean I didn’t have any other choices over how the labour and birth happened, but obviously the class has to cover a lot of different possibilities and it’s not going to cover every single one (Mum, 01)  ‘No, not at all, I’m afraid not at all. I had a lot of things to say about it though which would have been helpful, but it wouldn’t have prepared me for that… it didn’t really cover c-section.’ (Mum, 24) |
| **Missing information** | ‘Maternity bag, there was nothing about it. Practical stuff, just because it make me anxious arriving and not having everything I need, not going to the right place.’ (Mum, 07).  ‘There were some things that I came away and I didn’t really know too much about. I didn’t really feel like I knew enough about what induction was, and it was only when I did that in NCT a few weeks later that I found out really what it was.’ (Mum, 10)  ‘Well, it’s interesting cos at NCT we learnt about the pros and cons about different pain relief, such as pethidine and at the end of it most of us came away thinking we don’t want any of that, but when I speak to midwives it’s not an entirely different story, my midwife that I saw last week said maybe pethidine is something you’d like to consider it can be quite safe. It was almost at the ACE study as well they were saying that might be a good one to use. I felt sort of torn really between being given these facts and figures from the NCT and then it was slightly different hearing it from the midwife. And obviously the midwives are who I trust really. But when you get sort of objective facts and figures, so I still feel quite confused about pain relief options.’  ‘Perhaps guidance on writing a birth plan, we weren’t told much about that. How in detail it needs to be. Will it read and understood, will it be followed…. I’ve heard sometimes that it’s not.’(Mum, 010)  ‘The one thing it didn’t really cover… was the recovery stuff and what to expect from that.’ (Mum, 12)  ‘The reason I would attend the other classes is for the after [postnatal] stuff.’ (Mum, 12)  ‘Some of the techniques like the breathing techniques, I think it would be better to have a practice example, I don’t know if it was originally part of the curriculum or not.’ (Mum, 11)  ‘I would be interested to know the points where to press, lower back, upper back.’ (Dad, 11)  ‘At the beginning it mentioned different places where you can give birth like the [Midwife Led Unit] I didn’t quite understand the difference between those. Probably it’s obvious for native English people but for me it wasn’t.’ (Dad, 11) | A bit more about recovering immediately post the birth, I can’t remember if it was covered in all honesty (Mum, 03)  ‘I think the biggest thing we had was the stuff we had with us… you get these lists of these websites, but actually being really clear with what you needed would have been helpful.’ (Mum, 05)  ‘There was a birth preferences target left for us, which I thought was quite good but I’d already done my birth preferences by that point, but it was good way of looking at it’ (Mum, 05).  ‘It did go through induction and the stages of labour, but it didn’t really talk about the knock on effect of each thing might be, so if you did, say, take a pessary, then it would mean you weren’t able to have your baby at home. It didn’t talk about what each decision could make.’ (Mum, 24)  ‘Because it was a short course, it could have been a really good chance of signposting people to read certain books.’ (Mum, 24)  ‘More interactive stuff… interact with the resources, bits that were cut out, move around, things like that.’ (Mum, 24) |
| **Videos** | I think some of the video clips were quite helpful, just hearing, like, a real life experience (Mum, 01).  ‘mmm, I dunno, I’ve got so many stories from friends…. not that much.’(Mum, 07)  ‘Sort of useful I suppose, but umm didn’t go into too much detail so I didn’t feel I got too much from them.’ (Mum, 10)  ‘The projector wasn’t working so we just watched them off the laptop, but that was fine, there weren’t too many of us in the class really so we could see it.’(Mum, 10)  ‘…you know what might go wrong, what might not be what you expect, and it’s just helpful to hear from other people, how they felt.’ (Mum, 12)  ‘The videos didn’t make much difference, it’s not a bad thing but to me it added no value than a TV station.’ (Dad, 11)  ‘I think one of the videos the person was giving birth for the second time, so kind of knew what to expect.’ (Mum, 11) | ‘The mums’ stories were quite interesting, I quite liked those.’ (Dad, 01)  ‘I do remember the videos at the end, that was quite nice to hear others experiences.’ (Mum, 24) |
| **Asking questions** | ‘There was no such thing as a stupid question.’ (Mum, 02)  ‘We were encouraged to ask questions. I think the group size was quite nice because we felt comfortable to ask questions.’ (Mum, 10)  ‘Yeah, because I was able to ask questions specifically about what to expect at [hospital] for a c-section’ (Mum, 12)  ‘It was very easy to ask questions and the questions were well answered.’ (Mum, 11)  ‘There was one lady who kept asking questions, and I really liked that!’ (Mum, 13) | ‘I think I would have appreciated that [opportunity] to ask questions’.  ‘I felt like I could ask something, it felt like an approachable situation.’ (Mum, 24). |
| **Overall** | ‘I loved it, because I’m a healthcare practitioner, it was speaking to me quite well, because it was quite specific, with medical vocabulary. It was reassuring.’ (Mum, 07)  ‘I thought it was good, it was quite interesting because I’ve done a fair bit of hynobirthing preparation, some of the tone and the language was quite different to what I’ve been getting into my own headspace… was a little bit ‘oh’… but I thought some of the language the midwife used was quite interesting, and different from what I would have chosen…. It was quite important to feel empowered and like I’m going to be an important part of the labour and delivery.’ (Mum, 08)  ‘It kind of felt like it was matching the things I’d already heard about or was aware of.’ (Mum, 08)  We thought it was really good, we came away feeling really positive actually. We wondered if it would be a bit too similar to the NCT classes but it gave us quite a lot of extra in-depth information (Mum, 10)  ‘I actually thought it was really good. I’d already attended a six week antenatal class which covered stuff going beyond birth, and I found the ACE session was a lot more informative about what to expect leading up to labour and during labour (Mum, 12)  ‘I think the class was really well structured, it covered the relevant topics up to the delivery of the baby.’ (Mum, 11)  It made me understand more what [partner] is gonna go through. What I can expect, how can I help, it is much and not much but it helped me a lot’ (dad, 11)  ‘I have to say the presenter was very good.’ (Dad, 11)  ‘I thought it was brilliant, being pregnant at this time, it is quite lonely and stressful, so it’s quite nice there’s involvement.’ (Mum, 13)  ‘I found it very informative and for some reason I retained more information compared to the other class.’ The knowledge stayed with me’ (Mum, 13) | ‘It’s about getting the key information over in the time available and I think you did that really well’ (Mum, 003)  ‘There were definitely teething problems, the patter will become a bit more fluid as time goes on as the people who are presenting are getting used to doing the presenting.’ (Dad, 001)  ‘At one point the lady, the midwife said, you’ve got to listen to the doctor cos the doctor knows best, I felt like there was a real lack of trust in a woman’s body. And I didn’t feel like it covered enough, probably because it wasn’t long enough.’ (Mum, 24) |
| **Timings** | It was about right in length, maybe two sessions would be good. As one session it was a fine length of time, but maybe two sessions to cover any extra bits in a bit more detail. But it was good how it was quite concise’ (Mum, 10)  I think it was about right for what it covered if I’m honest. In fact I don’t think it lasted the whole 2.5 hours because by the end of that I was getting a bit fidgety. And I liked the pace of it, it didn’t feel too quick, because with the ones I’d been to before it felt a bit rushed.’ (Mum, 12)  I think it’s a bit too short, you can’t have enough information in two hours, but in a way it was enough because after two hours you get a bit tired… it’s not enough to icebreak, ask questions (Mum, 07)  Everything was concentrated in two hours, which is kind of into my style of gathering information, I like concentrated information, rather than let’s talk about one topic and then talk in groups.’ (Dad, 11)  ‘It was hard to keep sitting, maybe a little break, even though we were encouraged to take a break move around, you don’t want to miss information. Maybe a structured break?’ (Mum, 13) | ‘We expected it to be 2.5 hours, but my lift arrived at 2 and a quarter hours but because it overran a little bit… I left before it ended after 2h and 45 mins’  ‘It was very rushed, the two hours weren’t long enough.’ (Mum, 24). |
| **Resources** | [Posters] ‘A bit too small, I couldn’t see.’ (Mum, 07)  The pain relief options, that was really good, and then any sort of interventions like forceps or ventouse, that was good. We learned about that in probably more detail than we did in NCT all the sort of techniques and equipment, and also during a caesarean and how many people are there. Um, yeah, so that was good. And just to know that’s what can happen at [hospital] specifically. That was good to know from them (mum, 10)  There were diagrams and things dotted around the room which were quite useful, showing the stages of labour and the intensity of contractions and when they come and that was useful.’ (Mum, 10)  ‘I thought they were helpful, they were helpful particularly otherwise it’s just one person talking at you for 2 hours. She was really engaging, it just breaks it up a bit.’ (Mum, 12)  The assisted delivery methods… like the suction cup, I don’t know exactly, that was nice, I thought it was bigger, I thought it was like a plunger style, I don’t know why. We saw how small it was, I thought that was brilliant as well, you can easily manage to visualise what it would be like (Mum, 11).  ‘Also the visual representation with the balloon, for me it was like woah really, this is what’s really happening, it was very practical and very easy to understand and visualise what to expect… that balloon was like a thousand pictures.’ (Dad, 11) | Some of the illustration I thought was really quite nice, you’d had these pictures drawn by someone for, you know, to show that journey, I quite liked that, I’m quite a visual person. So that actual illustration and these are the things that, this is what a TENS machine looks like, these are the pain relieving options, I think those are really quite valuable for someone like me that thinks quite visually. Because obviously during the birth, the birthing process, it’s quite easy as the partner to be quite like swamped by what’s going on.. a lot of the stuff will go straight out of your mind… sometimes things that stick really well, something really simple to hang the bit of information on so when the time comes, I’m supposed to be doing this to help at this particular time’. (Dad, 01)  ‘It didn’t feel very well prepared, the resources didn’t necessarily match up to what was being talked about.’ (Mum, 24).  ‘There was a worksheet with different like circles on it, but we didn’t even look at it.’ (Mum, 24).  ‘I remember the pictures on the wall, very small, quite far away, difficult to look at.’ (Mum, 24) |
| **Social opportunities** | ‘There was no point, I didn’t interact with people (Mum, 007)  ‘But we just had more time to introduce ourselves and partners to introduce, it’s just not as awkward as asking questions… I think we introduced ourselves but only the ladies.’ (Mum, 13)  ‘..yeah, because there were some people, we had to social distance, but when you can see everyone’s faces, you feel like there’s people behind… it’s psychological isn’t it.’ (Mum, 13) | ‘Obviously looking in the context of the COVID situation at the time everyone was spaced out a little bit, so that sort of more natural perhaps opportunity to mingle and sort of interact with some of the Dads might have been nice.’ (Dad, 01) |
