## Supplementary file 5 for "Co-design and refinement of an optimised antenatal education session to better inform women and prepare them for labour and birth"

| Midwifery immediate feedback to researcher (general themes or important points raised by single person) | |
| --- | --- |
| Did the session go to plan | Question was answered in 12/19 sessions.  In 11/19 the feeling was yes but they altered the structure as required. 1/12 sessions they felt there wasn’t enough time to impart all the information. |
| Things that went well | Facilitators liked the structure  They liked it when groups were interactive  They felt that women engaged  There were positive comments on group size, especially when the size was smaller. |
| Could go better | Want more time so sections less rushed  Practical issues – IT not working well and poster falling down  Would like to increase interaction |
| Suggested improvements to session | Need more time (possibility to split into more sessions)  Need to add in more about emergency situations  Improve the way the resources and the way they work (e.g. poster peeling off the wall, better sound for the videos, bigger posters without American spellings, and make posters more professional)  Suggestions about integration of the coping strategies into the birthing stages  The order felt obstetric, need more early labour coping strategies (1 participant)  Two people presenting would be better |
| Feedback from midwifery focus group | |
| General | Not enough time would like longer session  Equipment didn’t work well at the start, others didn’t have a problem with the IT  Paid closer attention to births other than normal (not usual for ANE classes)  Initially emergency situations not included enough this change was made and it was good to include as usually not addressed  Should include more postnatal information – others encouraged attendance at local infant feeding session  ACE resources make it more interactive  Not sure that the birth preferences tool worked well as redesigned for covid. |
| Resources | Journey a good resource  Video concept is good but lacks diversity  Having resources e.g. pelvis and ping pong balls, kiwi  Would have liked forceps |
| Manual and training | Used it for checking up, needs some elements e.g. emergencies adding to it  Coping strategies needs to be interspersed throughout  The training was fine, but need an hour to prepare and then session evolves over time |
| Social opportunity and how to improve | Didn’t work well due to lack of interaction because of covid  Could introduce an interactive exercise for packing bag game  Could give out children’s centre registration form |
